# Combining Genetic Proxies of Drug Targets and Time-to-event analyses From Longitudinal Observational Data To Identify Target Patient Populations

**DOI:** 10.1101/2024.06.05.24308448

**Authors:** Luke Zhang, Prachi Kulkarni, Farshad Farshidfar, Whit Tingley, Tim Hoey, Whedy Wang, James R. Priest, Sylwia M. Figarska

## Abstract

**Background:** Human genetics is an important tool for identifying genes as potential drug targets, and the extensive genetic study of cardiovascular disease provides an opportunity to leverage genetics to match specific patient populations to specific drug targets to improve prioritization of patient selection for clinical studies.

**Methods:** We selected well described genetic variants in the region of *PCSK9* (rs11591147 and rs562556), *ADRB1* (rs7076938), *ACE* (rs4968782 and rs4363) and *BAG3* (rs2234962) for use as proxies for the effects of drugs. Time-to-event analyses were utilized to evaluate their effects on atrial fibrillation (AF) and heart failure (HF) death and/or re-hospitalization using real-world longitudinal dataset. To mitigate the effect of confounding factors for cardiovascular (CV) outcomes, we employed propensity score matching.

**Results:** After matching, a genetic proxy for PCSK9 inhibition (rs11591147) improved survival from CV death/heart transplant in individuals following a diagnosis of ischemic heart disease (Hazard Ratio (HR) 0.78, *P*=0.04). A genetic proxy for beta-blockade (rs7076938) improved freedom from rehospitalization or death in individuals with AF (HR 0.92, *P*=0.001), and a genetic proxy of ACE inhibition (rs7076938) improved freedom from death or rehospitalization for HF (HR 0.84, *P*=0.017) and AF (HR 0.85, *P*=0.0014). A protective variant in BAG3 (rs2234962) showed decreased risk of HF rehospitalization or CV death/heart transplant composite outcome within 10 years in HF patients (HR=0.96, *P*=0.033). Notably, despite smaller cohort sizes after matching, we often observed numerically smaller HRs and reduced P, indicating more pronounced effects and increased statistical association. However, not all genetic proxies replicated known treatment effects.

**Conclusions:** Genetic proxies for well-known drugs corroborate findings from clinical trials in cardiovascular disease. Our results may demonstrate a novel analytical approach that leverages genetic evidence from a large cohort to effectively select patient populations where specific drug targets may be most effective.

## Introduction

Drug discovery and clinical development is a high-risk process that may require decades of capital investment prior to clinical use with an estimated median cost of $1.1billion for each new drug approved^1^. Cardiovascular diseases with high heterogeneity such as heart failure (HF) frequently require complex and lengthy clinical trials enrolling thousands of patients to demonstrate efficacy^2^. However in some examples, a small effect in a large group may be derived from a large affect in a smaller subgroup of affected individuals ^3,4^. This phenomenon has been observed in a number of cardiovascular therapies, which have shown greater efficacy or clinical utility for specific subgroups of disease following regulatory approval^5,6^. Recognizing these nuances can guide the development of personalized medicine strategies, facilitating improved treatment effects in more targeted populations. This approach not only significantly reduces development costs but also optimizes therapeutic outcomes, thereby increasing the likelihood of success in drug development.

Regulatory bodies have issued guidance intended to increase the efficiency of drug development and support precision medicine by identifying subgroups of affected individuals where a specific therapeutic approach may be most effective^7^. Oncology has been at forefront of incorporating genetic information into drug discovery by identifying genetic targets which define prognosis in subtypes of large heterogeneous disease entities, and then designing therapies to specifically manipulate these targets^8,9^. Outside of oncology, human genetics has proven to be a useful tool for selection of drug targets, and may also have the potential to enhance the design and conduct of clinical trials in cardiovascular medicine.

Here we describe efforts to develop a novel analytical framework intended to generate a strong pre-clinical hypothesis about the patient population(s) where a therapeutic intervention is most likely to be effective using a real-world clinical dataset and time-to-event analyses. To test and validate our analytical approach for cardiovascular disease, we leverage genetic variants with a known biological effect on gain, loss of function, or protein expression levels, as well as expected associations with cardiovascular phenotypes. With such variants we construct *in silico* trials using real world data and compare the results to the outcomes of well-established randomized clinical trials of approved therapies. We demonstrate the potential utility of our analytical framework using known therapeutic targets across diverse areas of cardiovascular medicine, including lipid biology to atherosclerosis outcomes, beta-adrenergic signaling to HF and atrial fibrillation (AF) outcomes. Furthermore, we aim to explore whether a therapy originally developed for treating Mendelian cardiomyopathy, a specific genetic heart condition, might also offer therapeutics benefits to a broader population of individuals diagnosed with HF.

## METHODS

### Study Population, Clinical Data, and Outcomes

Genetic and clinical data in the UKB cohort were obtained from the UKB (https://www.ukbiobank.ac.uk) and is available to researchers through a streamlined application process. The UKB was approved by the North West Multi-Centre Research Ethics Committee and all participants provided written informed consent to participate in the study. We used International Classification of Diseases ICD-9 and ICD-10 codes to define patients’ populations, co-morbidities and cardiovascular outcomes. Detailed ICD codes are provided in Supplemental Table 1. We considered two outcomes for this study: a) CV death or heart transplant (CV death/heart transplant), and b) rehospitalization or CV death/heart transplant - composite outcome. Details on selection of covariates (co-morbidities and medication use) considered for propensity score matching for the specific outcome are described in the supplementary material.

### Statistical Approach

All statistical analyses were performed in R (version 4.3.3) using the *MatchIt* for propensity score matching, and *survival* packages for time-to-event analyses. For inclusion, exclusion, and matching of clinical covariates which can vary with time (such as medication usage, pre-existing conditions), individuals were matched based on covariates at the time of the initial diagnosis. For each genetic variant, individuals with effect allele were defined by variant allele (heterozygous or homozygous) and compared to individuals with the other allele. Covariate balance post-matching was assessed using standardized mean differences in Love plots, generated with the love.plot() function in *cobalt* R package, examining the balance measures before and after conditioning. Survival analyses were performed using Cox Proportional Hazards model and visualized with survplot as Kaplan-Meier or Cox Hazards curves as appropriate. For each variant analyzed, we also performed a post-hoc power calculation based upon the available population size, number of events, and follow-up time, using the *SurvSNP* R package (details are provided in the Supplementary Material). The goal was to enhance our understanding of the reliability of our findings and to potentially guide future research endeavors. Additionally, for each SNP we conducted a PheWAS using PHEnome Scan ANalysis Tool (PHESANT)^10^ and details of the analysis are provided in the Supplementary Material.

### Genetic Variants

Similar to the approach for instrumental variable selection in Mendelian Randomization, we selected a set of genetic variants with clearly explainable biological impact on a gene which is the target of either an approved or proposed therapeutic modality. A missense variant in *PCSK9* (rs11591147 p.Arg46Leu) represents the effect of PCSK9 inhibition and has a protective effect on hyperlipidemia and coronary heart disease^11^. We also tested another *PCSK9* missense variant, (rs562556 p.Val474Ile) that is likely to have an impact on PCSK9 activity and previously showed associations with LDL levels and CAD risk^11,12^. Another variant of interest is rs7076938 located upstream of beta-adrenergic receptor beta-1 (*ADRB1)* (tagging for Gly389Arg rs1801253 r^2^=0.96) that increases blood pressure and thus mimics effects of beta-adrenergic agonists. A genetic variant, rs4968782, located upstream of the Angiotensin-converting enzyme gene (*ACE*) likely affects transcription factor binding, and is a strong eQTL associated with increased *ACE* expression in the lung; therefore mimicking the effect of ACE inhibitors to modulate risk of hypertension. Also, effects of this SNP are stronger than effects of coding variants that were initially linked to circulating ACE levels and ACE activity^13^. We also tested an intronic variant in *ACE*, rs4363, that previously showed a very strong association^14^ with ACE levels reaching significance of *P*=2×10^-257^. This intronic SNP is highly correlated (r^2^=0.9 in EUR) with a missense variant rs4343 associated with ACE activity^15^. While no therapy is currently approved for *BAG3*, a coding variant in *BAG3* gene (C151R) is associated with decreased incidence of HF and has been mechanistically demonstrated to increase the protective effect of the BAG3 protein from proteo-toxicity in cardiomyocytes^16^.

## Results

As a first pass diagnostic analysis, we aimed to replicate the previously described case-control associations with the selected variants in the UKB, all of which showed associations with expected phenotypes (Supplemental Table 2). The numbers of allele carriers per variant and event are presented in the figure panels (Figures 3-6) corresponding to each SNP. For patients who did not experience an event, follow-up period was calculated from the time of disease diagnosis until the most recent data update from the UKB, version September 2023. Events of non-cardiovascular death were right censored at the time of death. To evaluate the composite outcome of rehospitalization for disease of interest (HF or AF) or CV death/heart transplant, the follow-up period from the time of diagnosis was censored at the number of years where the impact of the SNP was most representative. All the tested variants met the underlying assumptions for proportional hazard risk over time as indicated by log-log plots.

The analytical framework (Figure 1) consists of selecting the individuals heterozygous or homozygous for the alternative allele for the variant of interest, selection of relevant known clinical risk factors including age at primary diagnosis, clinical comorbidities, and medication use. Cox Proportional Hazard models were performed prior to matching (with adjustments for age at disease diagnosis and sex), and after matching (without adjustments). Given the known associations for each of the selected variants and the intent of replicating known therapeutic effects, for the purposes of the methods development we defined statistical significance as a nominal *P* less than 0.05 and we did not perform correction for testing of multiple hypotheses. When the number of individuals homozygous for the effect allele was less than 25% of the number of heterozygous individuals, the groups were combined into a single group after matching. For each variant analyzed, we also performed a post-hoc power calculation based upon the available population size, number of events, and follow-up time. A summary of start and end timepoints for survival analyses conducted for each SNP, as well as the number of patients included in the study, can be seen in Figure 2. Additionally, power calculations for survival analyses conducted for each SNP can be seen in Supplemental Table 3.

**Figure 1.**
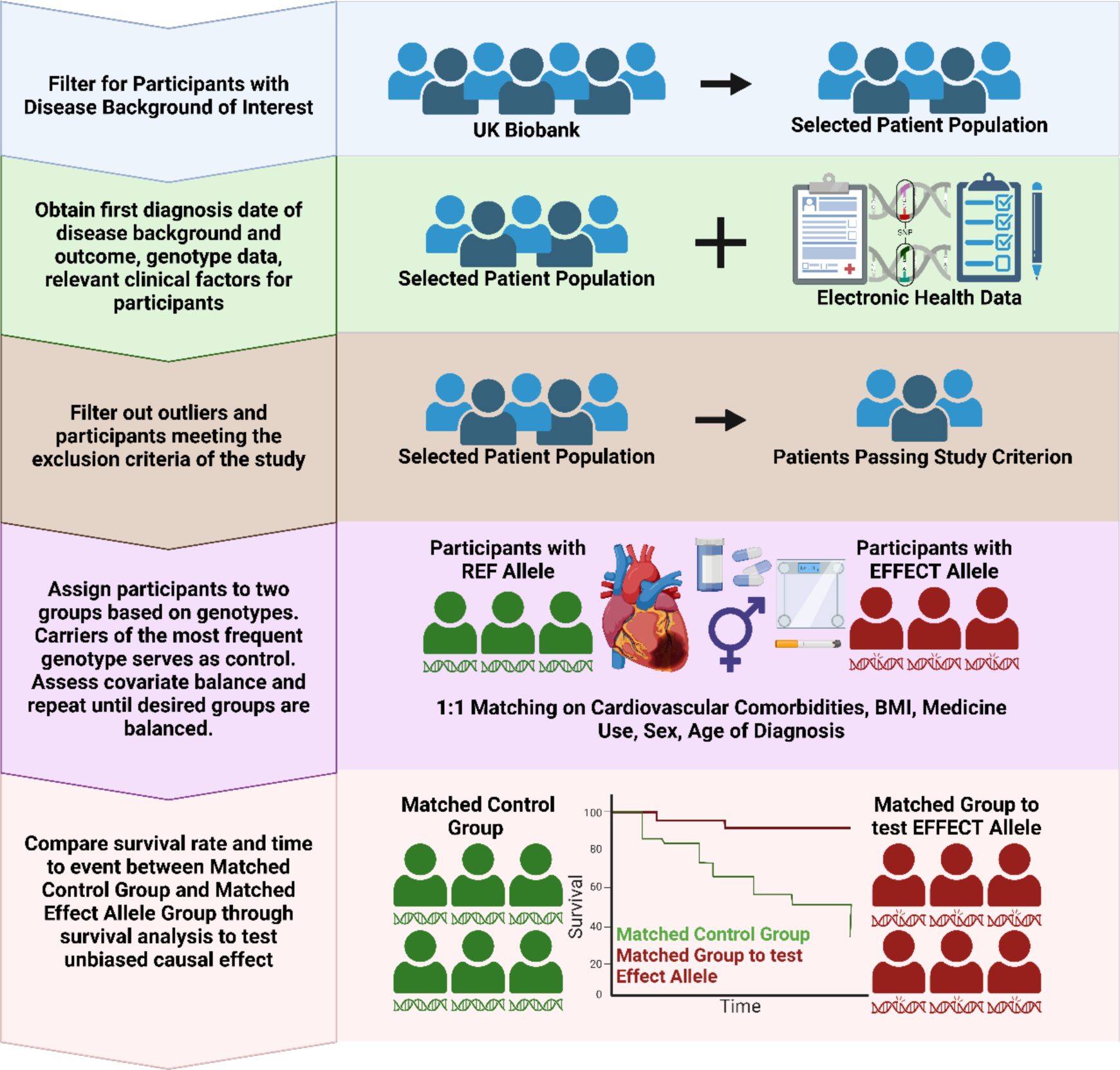
Analytical framework. **Participants diagnosed with a disease of interest were included in the study. Information on co-morbidities and medication use was obtained and used for propensity score matching procedure. Survival analysis was performed for matched allele carriers.**

**Figure 2.**
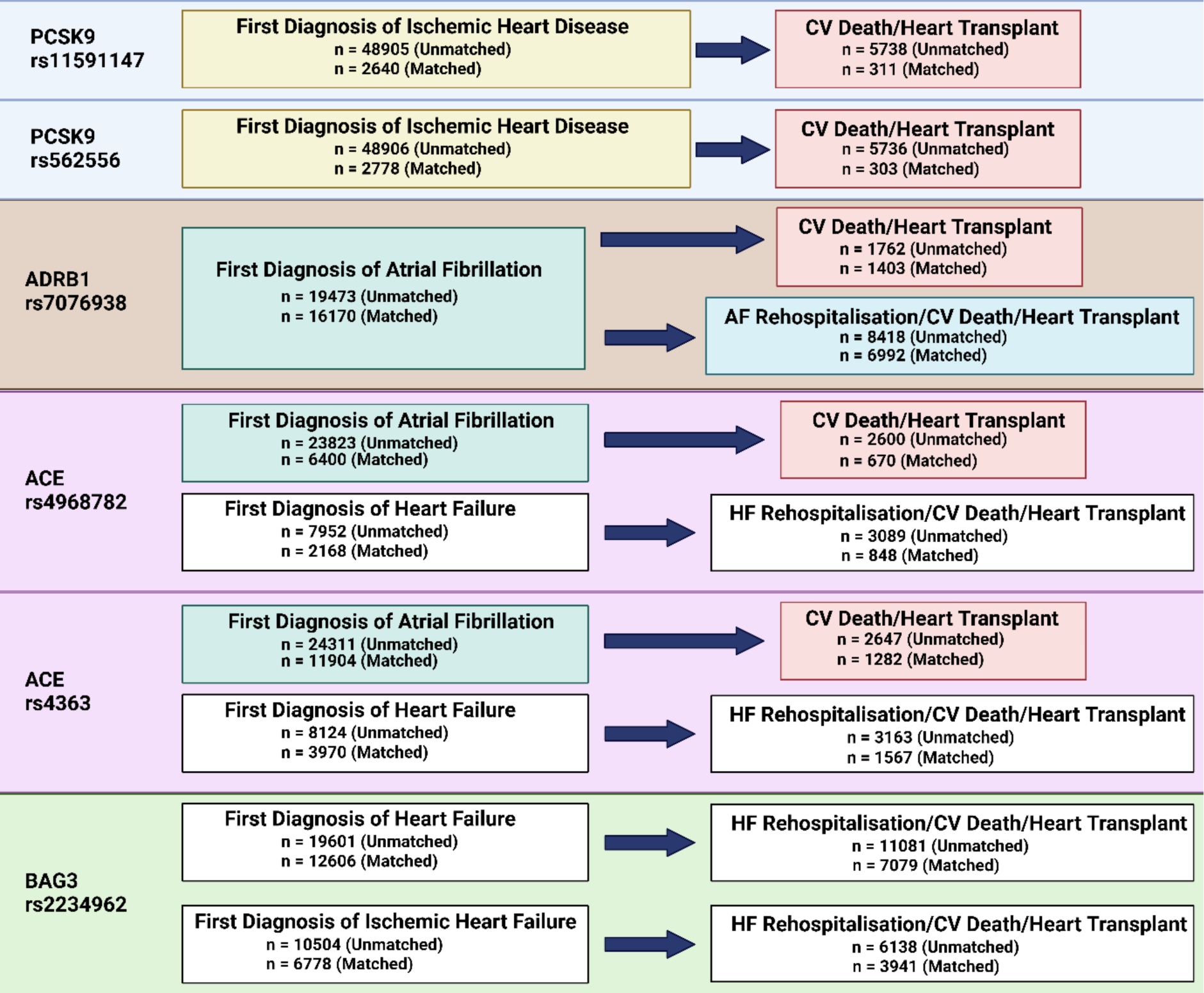
Summary of all survival analysis performed for each SNP in the study, indicating disease of interest and considered outcomes.

### PCSK9 rs11591147

PCSK9 is a well validated genetic target with a clearly defined mechanism of action to reduce circulating cholesterol levels and multiple successful clinical trials for prevention of atherosclerotic cardiovascular disease (ASCVD). The LoF variant rs11591147 disrupts the function of the *PCSK9* which results in higher amount of cell surface LDLR and internalization of LDL, leading to lower serum LDL levels and subsequently confer protection against ischemic heart disease (IHD)^17,18^. To replicate the successful trials for PSCK9 inhibitors for secondary prevention of ASCVD, we examined cardiovascular death or heart transplant among individuals after a diagnosis of IHD. The impact of rs11591147 on time to CV death/transplant among genetically determined Caucasians diagnosed with IHD did not reach statistical significance, exhibiting a hazard ratio (HR) of 0.95 and a *P* of 0.53. However, after matching on comorbidities existing at the time of diagnosis of ASCVD (Supplemental Figure 1), medication usage, and cardiovascular comorbidities, carriers of one or more effect alleles (T) demonstrated improved survival compared to individuals with no copies of the effect allele (Figure 3a). The HR markedly decreased to 0.78 with a significant *P* of 0.03, indicating a protective effect of PCSK9 rs11591147 against the development of severe cardiovascular outcomes from an IHD diagnosis. The difference between the unmatched and matched HRs underscore the importance of accounting for other causal factors, including some that were implemented as part of a treatment strategy, in determining the true impact of PCSK9 inhibition on cardiovascular outcomes (Figure 3). Results of covariate balancing for each matching term are presented in Supplemental Figure 1.

**Figure 3.**
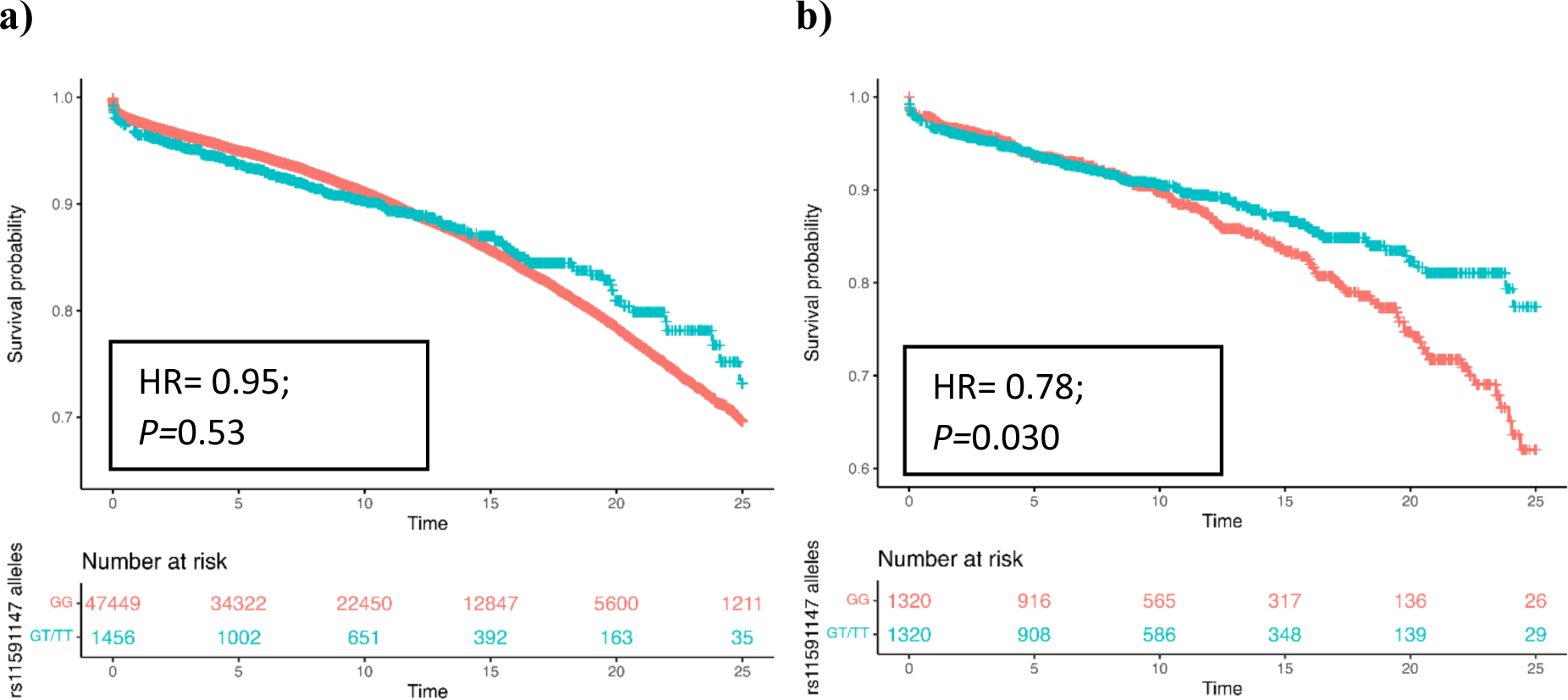
Effect of PCSK9 rs11591147 on time to cardiovascular (CV) death/heart transplant from first ischemic heart disease diagnosis (IHD) in a) unmatched and b) matched participants. Only Caucasians were included. Individuals with GT/TT genotypes (proxy for PCSK9 inhibitor treatment) showed better survival for CV death/heart transplant compared to individuals with GG genotype (controls).

### PCSK9 rs562556

The impact of rs562556 on time to CV death/transplant from date of first IHD diagnosis among Caucasians did not reach statistical significance, exhibiting a HR of 1.01 and *P* of 0.342. Even after matching on comorbidities (Supplemental Figure 6) existing at the time of diagnosis of ASCVD, medication usage, and cardiovascular comorbidities, no statistically significant impact was observed by rs562556 (HR=1.06, *P*=0.641, Supplemental Figure 5).

### ADRB1 rs7076938

ADRB1 encodes the β_1_ adrenergic receptor with primary cardiovascular effects in the heart sinus node and juxtaglomerular cells of the kidney to modulate contractility, heart rate, and blood pressure. It is the target of many selective beta blockers used in primary and secondary prevention of ASCVD, HF, hypertension, and AF^19^. The variant rs7076938 is a tagging SNP for rs1801253 (Gly389Arg, r^2^=0.96 in EUR and r2=0.81 in all populations) that has been shown to alter post-receptor signaling, with the Arg389 receptor (tagged by rs7076938 T) coupling more efficiently with its corresponding G protein (Gs)^20–22^ and correlating with higher blood pressure^23,24^. In our study we tested rs7076938, as it was more significantly associated with SBP in large genome wide association studies (GWAS)^24^ and was used as a part of instrument variable in MR analysis representing antihypertensive medication proxy^25^. Analogous to successful trials of beta blockers for treatment of AF, we examined AF progression with the combined outcome of rehospitalization for AF or CV death/heart transplant^26^. Carriers of TT genotype had increased risk of CV death/heart transplant since first AF diagnosis (HR=1.169, *P*=0.0012) and the effect remained significant in the matched data (HR=1.172, *P*=0.0031). Additionally, carriers of the TT genotype showed increased risk of AF rehospitalization or CV death/heart transplant in unmatched (HR=1.01, *P*=0.0043) and matched participants (HR=1.081, *P*=0.0012). The impact of ADRB1 on cardiovascular outcomes before and after matching can be seen in Figure 4. Results of covariate balancing can be seen in Supplemental Figure 2.

**Figure 4.**
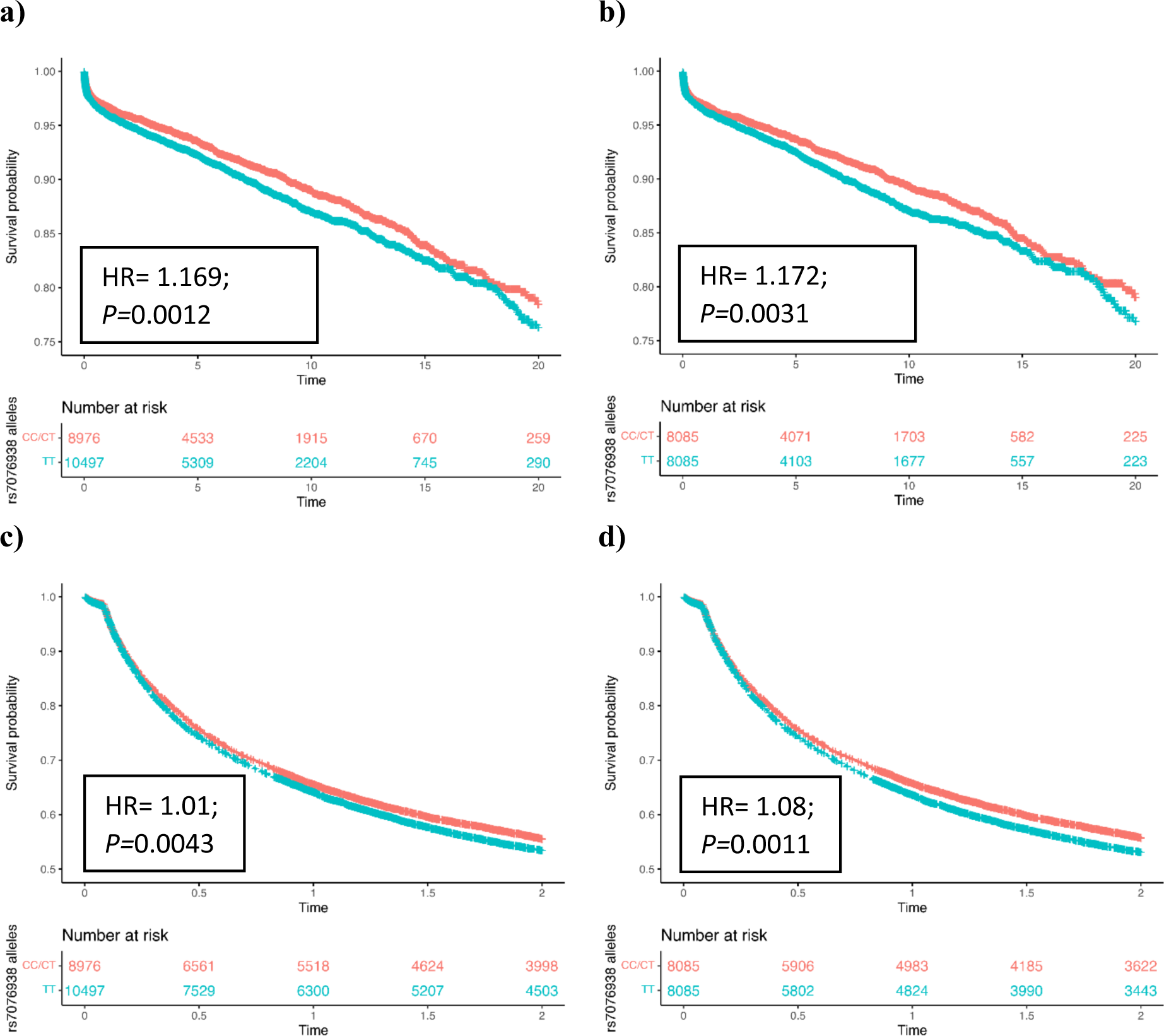
Effect of ADRB1 rs7076938 on time to cardiovascular (CV) death/heart transplant from first atrial fibrillation (AF) diagnosis in a) unmatched and b) matched participants, and time to AF rehospitalization or CV death/heart transplant in c) unmatched and d) matched participants. Individuals with TT genotype showed increased risk of AF rehospitalization of CV death/heart transplant composite outcome compared to individuals with CC/CT genotypes.

### ACE rs4968782

The ACE gene encodes the angiotensin converting enzyme expressed in the lungs which is a key step in the renin/aldosterone-angiotensin system regulating vascular tone and blood pressure. The variant rs4968782 likely affects transcription factor binding and is a strong eQTL affecting ACE levels. Inhibitors of ACE such as lisinopril are a mainstay of treatment for hypertension, HF, and AF. Carriers of at least one A allele exhibited decreased risk of CV death/heart transplant from first AF diagnosis in unmatched (HR=0.94, *P*=0.029) and matched (HR=0.85, *P*=0.0014) participants. When the composite outcome – HF rehospitalization or CV death/heart transplant – was considered, the impact of the A allele was only seen in matched data (HR=0.84, *P*=0.017 vs HR=0.93, *P*=0.14 in unmatched data). It is important to note that the time-to-event analysis of CV death/heart transplant from first AF diagnosis was performed in the Caucasians as the SNP only showed effects in this subset, while time to composite outcome was performed in the entire population regardless of ethnicity. These results underscore the impact of propensity score matching in elucidating the significance of the ACE SNP in cardiovascular outcomes, revealing previously undetected associations, and enhancing the precision of risk assessments (Figure 5). Results of covariate balancing are displayed in Supplemental Figure 3.

**Figure 5.**
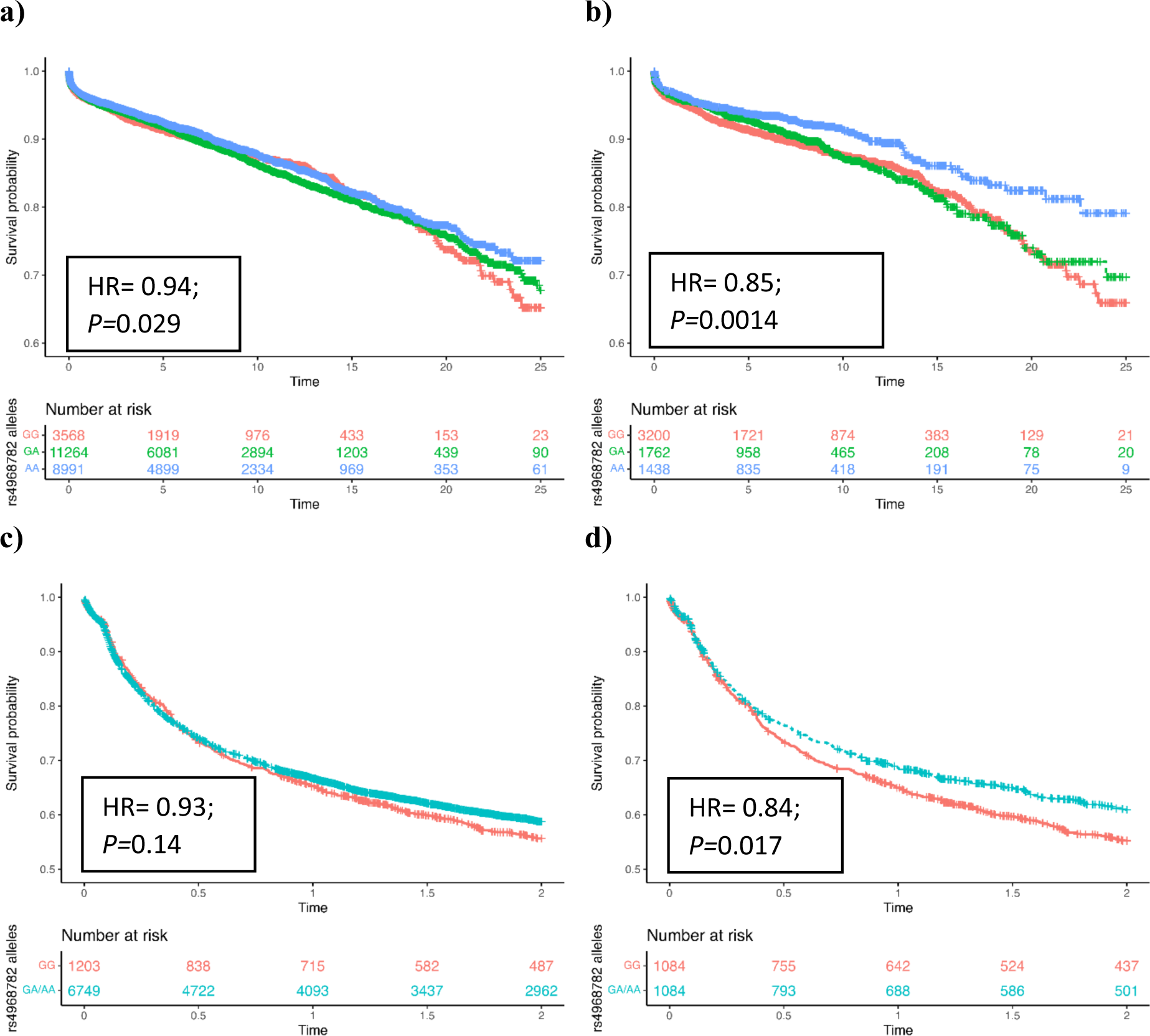
Effect of ACE rs4968782 on time to cardiovascular (CV) death/heart transplant from first atrial fibrillation (AF) diagnosis in a) unmatched and b) matched Caucasians, and time to heart failure (HF) rehospitalization or CV death/heart transplant in c) unmatched and d) matched participants of any ethnicity. Carriers of GA or AA genotype showed better survival for rehospitalization and severe cardiovascular outcome compared to individuals with GG genotype.

### ACE rs4363

For ACE rs4363, carriers of the A allele did not exhibit decreased risk of CV death/heart transplant from first AF diagnosis (HR=0.97, *P*=0.197 in unmatched, HR=0.95, P=0.17 in matched). When the composite outcome – HF rehospitalization or CV death/heart transplant was considered, the impact of the A allele was not seen in either unmatched (HR=1.01, P=0.89) or matched data (HR=1.04, P=0.45). It is important to note that the time-to-event analysis of CV death/heart transplant from first AF diagnosis was performed in Caucasians, while time to composite outcome was performed in the entire population, regardless of ethnicity in order to mirror the analysis of ACE rs4968782. Survival analysis results can be seen in Supplemental Figure 7, while the results of covariate balancing can be seen in Supplemental Figure 8.

### BAG3 rs2234962 (C151R)

BAG3 plays an important role in the protein quality control pathway within cardiomyocytes, and loss of function mutations can cause a form of dilated cardiomyopathy^28^. The protective effect of the C allele on the composite outcome of HF rehospitalization or CV death/transplant within 10 years was observed in all HF patients (HR=0.96, *P*= 0.033) and remained essentially unchanged after matching (HR=0.95, *P*=0.011). Since a recent study has shown that delivery of additional Bag3 to the heart using AAV9 may result in improved functional parameters in the myocardial infarction model of mouse HF^29^, we tested effect of the SNP in the subset of HF patients of ischemic origin. We observed a protective HR against HF rehospitalization or CV death/transplant composite outcome 10 years in matched data (HR=0.95); however, this effect did not reach statistical significance (*P*=0.059and HR=0.97, *P*=0.202 in ischemic HF patients before matching, Figure 6). Results of covariate balancing are presented in Supplemental Figure 4. There was no observed effect of rs2234962 on ischemic HF or all cause HF when CV death/heart transplant was considered independently (HR=0.99, *P*= 0.703 and HR=1.01, *P*= 0.803 respectively in unmatched data).

**Figure 6.**
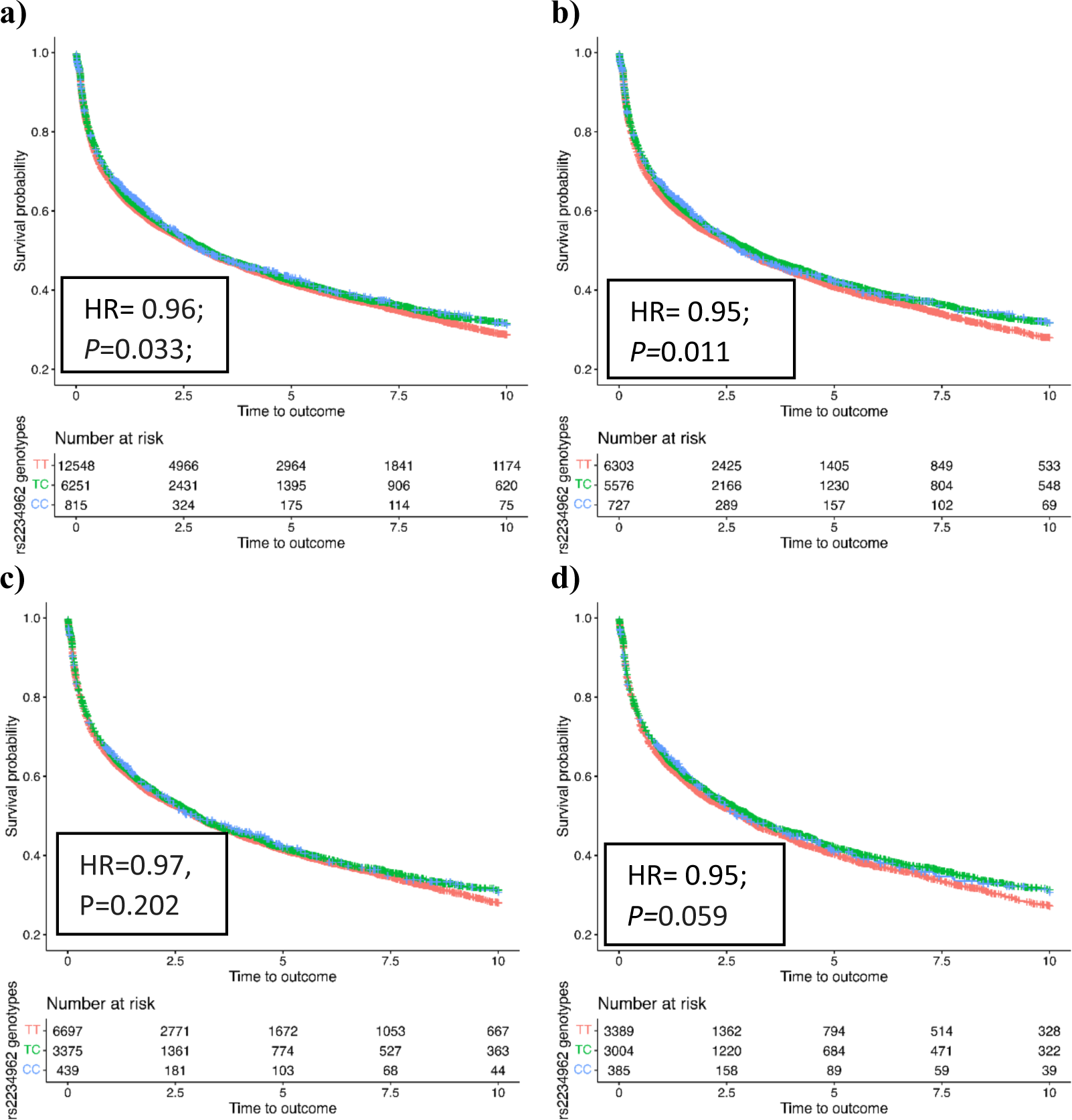
Effect of BAG3 rs2234962 on time to composite outcome – heart failure (HF) rehospitalization or cardiovascular (CV) death/heart transplant from first HF diagnosis in a) unmatched and b) matched participants, and on time to composite outcome – HF rehospitalization or CV death/heart transplant from first HF diagnosis due to ischemic origin in c) unmatched and d) matched participants. Carriers of the C allele showed decreased risk of HF rehospitalization or CV death/heart transplant composite outcome in HF patients compared to those with the T allele.

PheWAS was conducted for all the SNPs mentioned above, with the aim of finding associations mentioned in previous publications. PheWAS results for each individual SNP can be seen in Supplemental Figures 9-14.

## DISCUSSION

Here we describe a genetic survival analysis which employs a well-characterized genetic instrumental variable as a proxy for potential treatment with a specific drug or target. This approach has the potential to identify treatment effects using real-world outcome data through time-to-event analyses. In three examples of targets with approved therapies (PCSK9, ADRB1, ACE), this analytical framework identifies beneficial treatment effects seen in clinical trials, and a potential benefit in a clinical scenario for a novel target (BAG3) suggested by mouse models of ischemic cardiomyopathy. The approach described here is a natural extension of GWAS of clinical outcomes, Mendelian Randomization, and similar time-to-event GWAS^30–32^. Genetic survival analysis offers a critical advantage over standard applications of GWAS or Mendelian Randomization which typically only consider non-genetic covariates such as age and sex. Our proposed approach not only estimates the direction of effect for a specific drug or target in a real-world setting, but it also incorporates clinical covariates, causal factors, and standard of care therapies commonly encountered in clinical trials.

Matching is a well-accepted statistical technique to mitigate the influence of measured confounders and constitutes an essential part of clinical trial design often implemented in the form of ‘randomization’. Although matching on the propensity score is often effective at eliminating differences between the allele carrier groups to achieve covariate balance, the performance of this method must be carefully assessed. In addition to covariate balance, the quality of the match is determined by how many units remain after matching^33,34^. Matching often involves discarding units that are not paired with others. Moreover, some matching options, such as setting restrictions for common support or calipers, may further reduce the number of units available for analysis. In our research, successful matching often led to a notable reduction in sample size but also a more pronounced effect of the SNP. This suggests that our matching procedure effectively mitigated the influence of confounding factors and enabled us to discern a true effect of the SNP. Put differently, the improvement in power to detect known effects of genetic instruments for PCSK9, ADRB1, and ACE following matching underscores the methodological robustness of our approach and highlights its compatibility with complexities inherent in real-world longitudinal observational data. It is important to note that matching did not resolve all the complexities of real-world data, as the ACE variant rs4363 did not appear to modify the risk of adverse outcomes in AF or HF, which contradicts findings from previous clinical trials.

Cardiovascular diseases such as HF are frequently complex, where a broad diagnosis may arise from a combination of one and often more environmental and genetic causes. Findings from genetic data such as GWAS hold great potential in guiding drug discovery, as these studies have the unique capability for identifying risk loci for a disease without an *a priori* hypotheses. While GWAS is useful in identifying novel genetic loci that are beyond our current understanding of the disease, cell-based or animal models are unable to fully mimic the human condition, limiting the success rate of translating preclinical findings into clinical practice. Additionally, the genetic factors identified from GWAS for disease susceptibility may not always be the same factors that govern disease progression or long-term outcomes—which are often the endpoints of clinical trials and the most meaningful outcomes to affected individuals and physicians. Therefore, for drug target prioritization, more studies based on clinical samples of affected individuals with real world phenotype data may more accurately reflect the genetic basis of the condition under study. To ensure precise investigation of specific patient populations, our study utilized only Hospital Episode Statistics (HES) data and ICD codes, excluding self-reported cardiovascular complaints without direct medical diagnoses. Further, aligning with best practices in clinical trial enrollment, our analysis did not limit participants to Caucasian individuals —except in noted cases— and included all participants, irrespective of ethnicity.

Like other techniques based upon human genetics, the application and interpretability of genetic survival analysis has important limitations. Coding and non-coding common genetic variants that may be selected as instrumental variables typically have small effect sizes, which may be obscured by other clinical factors unless the number of individuals and events are relatively large. Conversely, the power to detect survival benefit may be compromised even with large effect sizes seen with rare variants which may suffer from a small number of individuals and events. Additionally, while our findings illustrate that matching on comorbidities and stratification appear to be useful tools for ‘focusing’ the technique to enhance the power, they do not eliminate the possibility of reverse causality or collider bias. Such biases can occur when individuals that carry a particular genetic variant have a measurably different risk of disease susceptibility —often observable in case-control GWAS — which impacts long-term disease outcome^35,36^. The analytical framework may only be applied when a genetic instrumental variable is present in the population of interest. An additional limitation of our current application is its inability to provide a meaningful estimate of treatment effect size for the target, which is among the most critical information derived from clinical trials for regulatory bodies, commercial entities, and payors^37,38^.

A significant strength of the genetic survival analysis work presented here lies in the diversity of genetic variants examined and the broad range of cardiovascular outcomes assessed, including IHD, HF, and AF. This comprehensive approach helps identify specific patient populations that would benefit most from potential treatments.

A logical extension of the analytical framework presented here would be to conduct *in silico* trials to select and optimize specific clinical variables and co-morbidities, with the goal of maximizing effect size (and minimizing the cost and duration of a clinical trial) for a new treatment. This approach of stratification might not be limited to clinical variables or co-morbidities but might also logically include other genetic factors such as polygenic scoring or stratification by Mendelian forms of cardiovascular disease. This leads to an important limitation of the analyses presented here; though we chose genetic instruments largely based on efficacious and widely accepted therapies, our findings are derived from a single large cohort study (UKB) and would benefit greatly from validation with an independent dataset.

## Conclusions

Our study’s findings suggest that genetic survival analysis could be instrumental in generating clinically meaningful hypotheses. This method identifies individuals and diseases that are likely to benefit from treatment of a specific target, and therefore could help guide the design of future clinical trials.

## Data Availability

Genetic and clinical data in the UKB cohort were obtained from the UKB (https://www.ukbiobank.ac.uk) and are available to any qualified researcher worldwide through a streamlined application process.

## Acknowledgments

This research was conducted using the UK Biobank Resource under Application number 84103.

## Sources of Funding

All authors are employees of Tenaya Therapeutics.

## Disclosures

All other authors declare no competing financial interests.

## Code and Examples

https://github.com/tenayatherapeutics/Genetic-Survival-Analysis-in-UKB/tree/main

## Supplementary Material

Supplemental Methods

Tables S1-S3

Figures S1-S14

## SUPPLEMENTAL MATERIAL

### Study samples

The UK Biobank (UKB) is a prospective cohort study with genotype and detailed phenotype information for up to 502,269 individuals aged 40-69 years when recruited between 2006 and 2010. UKB participant data, linked to NHS central registers for information on death, cancer registrations, and hospital admissions in England, Scotland, and Wales up to September 1^st^, 2023, are enriched with demographic characteristics, lifestyle details, and self-reported health outcomes. This comprehensive dataset, which includes hospital admission records from 1985 for Scotland and 1997 for England and Wales, undergoes record linkage based on NHS number, postcode, sex, and age. Collected baseline information encompasses various facets, such as age, gender, ethnicity, country of birth, education, socioeconomic status, BMI, and smoking status. Extracted from linked HES data are dates, duration of hospitalization, ICD-10 diagnostic codes, and OPCS-4 procedural codes. Genotype data is also available for most participants, which includes positions that are either directly genotyped on one of two arrays (UKB Axiom and UK BiLEVE arrays) or imputed using a reference panel from the Trans-Omics for Precision Medicine (TOPMed). Participants who did not have genetic data were excluded from this study. First available diagnosis dates, derived from HESIN (Hospital inpatient data) operations and diagnosis were considered for this study. Rehospitalization was defined as the next instance occurring 30 days after the first diagnosis, with endpoints including rehospitalization, cardiovascular death, study end (September 1^st^, 2023). Events of non-cardiovascular death were censored at the time of death. The starting dates correspond to when a patient was initially diagnosed with a specific disease of interest. UK Biobank was approved by the North West Multi-Centre Research Ethics Committee and all participants provided written informed consent to participate.

### Propensity score matching

Propensity score matching, implemented through nearest neighbor stratification using the *MatchIt* R package (Ho D, Imai K, King G, Stuart E (2011). “MatchIt: Nonparametric Preprocessing for Parametric Causal Inference.” Journal of Statistical Software, 42(8), 1–28. doi:10.18637/jss.v042.i08.), was a pivotal method employed to enhance comparability between treatment and control populations for the subsequent assessment of survival outcomes. The technique involved assigning individuals with available genetic data (either genotyped or TOPMed imputed) into control and treatment groups based on their reported SNP dosage, typically categorized as rounded dosages of 0 and those having at least one of the effect alleles (combining 1 and 2 dosages). Propensity scores were generated to represent the probability of receiving the genetic treatment, considering various baseline covariates.

To ensure a well-balanced comparison, participants were meticulously matched on critical factors listed for each SNP below. The evaluation of balance before and after matching was conducted using the standardized mean difference, with an absolute value less than 0.1 for total distance serving as a criterion for well-balanced groups. Propensity score matching not only facilitated the control of potential confounding variables but also enhanced the validity of subsequent survival analyses by creating more comparable cohorts based on clinical characteristics.

The study focused on testing single nucleotide polymorphisms (SNPs) associated with known cardiovascular effects, specifically examining rs11591147 and rs562556 from PCSK9, rs7076938 from ADRB1, and rs4968782 as well as rs4363 from the ACE gene. Regression analyses were conducted using glm function of the *glmnet* package in R, employing both Gaussian and binomial models linear and binary outcomes. The glmnet package (Version 4.1-8; Friedman et al., 2010) facilitated data analysis. Genotypic information was available for 464,428 participants for rs4968782, 473,376 participants for rs7076938, and 487,227 participants for rs11591147. This robust dataset enables a comprehensive exploration of the genetic associations between these selected SNPs and cardiovascular outcomes, providing valuable insights into the potential role of these genetic variations in cardiovascular health.

The rs11591147 and rs562556 polymorphisms within the PCSK9 gene were investigated, revealing an overall mean standardized difference of less than 0.01 in the examined cohort. The control group consisted of participants homozygous for the G allele while the treatment group consisted of participants with at least one copy of the T allele for rs11591147, or at least one copy of the A allele for rs562556. Covariate adjustments were implemented for sex, C-Reactive Protein, smoking, body mass index (BMI), low density lipoprotein (LDL) levels, first systolic blood pressure reading, diuretics use, angiotensin II receptor blocker (ARB) use, beta-blockers use, ACE inhibitor (ACEi) use, calcium channel blocker use, digoxin use, statins use, anti-coagulants use, age of ischemic heart disease (IHD) diagnosis, and previous disease diagnoses encompassing atrial fibrillation (AF), aortic valve stenosis, chronic renal failure, chronic obstructive pulmonary disease (COPD), heart failure (HF), dilated cardiomyopathy (DCM), hypertrophic cardiomyopathy (HCM), mitral valve disorder, type 1 diabetes (T1D), type 2 diabetes (T2D), unspecific stroke, and ventricular arrhythmias. The exclusion criteria for this study encompassed patients undergoing treatment with ezetimibe and centrally acting anti-hypertensive and vasodilator anti-hypertensive medications. These exclusions were instituted due to the pharmacological actions of these agents, which target LDL cholesterol, akin to the mechanism of PCSK9 inhibitors. In this analysis of PCSK9, only genetically determined Caucasian patients were included due to the observed heightened effect of the SNP within this demographic, enhancing detection sensitivity and statistical power for assessing its influence on the phenotype. Analysis in unmatched data was adjusted for sex and age of disease diagnosis, while analysis in matched data was unadjusted (Supplemental Figure 1).

The rs7076938 polymorphism within the ADRB1 gene was investigated, revealing an overall mean standardized difference of less than 0.01 in the studied population. The control group consisted of participants with at least one copy of the C allele, while the treatment group consisted of participants homozygous for the T allele. Covariate adjustments were made for various factors including sex, C-Reactive Protein level, smoking status, BMI, LDL levels, diuretics use, calcium channel blocker, alpha adrenoceptor blocking drug use, statins use, digoxin use, ARB use, anti-coagulant medication use, previous disease diagnosis for aortic valve stenosis, chronic renal failure, (COPD, HF, T2D, T1D, ischemic heart disease (IHD), mitral valve disorder, unspecific stroke, and age of AF diagnosis. We excluded individuals using ACEi or beta-blockers to mitigate confounding effects associated with the excluded medications (Supplemental Figure 2). This matching criterion was used for both the assessment of severe cardiovascular outcomes (CV death/heart transplant) after AF diagnosis and time to rehospitalization or CV death/heart transplant composite outcome from AF. Analysis in unmatched data was adjusted for sex and age of disease diagnosis, while analysis in matched data was unadjusted.

The rs4968782 and rs4363 polymorphisms within the ACE gene was scrutinized, revealing an overall mean standardized difference of less than 0.01 in the studied population. For each SNP, the control group consisted of participants homozygous for the G allele while the treatment group consisted of participants with at least one copy of the A allele. When investigating time to rehospitalization due to all-cause HF or CV death/heart transplant in patients diagnosed with HF, we balanced for sex, C-Reactive Protein level, smoking status, BMI, LDL, diuretics use, calcium channel blocker use, statins use, digoxin use, ARB use, anti-coagulant medication use, aortic valve stenosis status, chronic renal failure status, COPD status, HF status, T2D status, unspecific stroke status, IHD status, and age of HF diagnosis. We excluded individuals using ACEi, beta-blockers, alpha-adrenoceptor blocking drugs, vasodilator anti-hypertensives, and centrally acting anti-hypertensives to mitigate potential confounding effects associated with these medications (Supplemental Figure 3).

Furthermore, in the assessment of severe cardiovascular outcomes (CV death/heart transplant) after first available AF diagnosis, we balanced the data for sex, C-Reactive Protein, smoking, BMI, LDL, diuretics, beta blockers, calcium channel blockers, alpha-adrenoceptor blocking drugs, statins, anticoagulants, diabetic medications, age of AF disease diagnosis, and previous disease diagnosis for aortic valve stenosis, chronic renal failure, COPD, HF, unspecific stroke, IHD, T2D, unspecific stroke, ventricular arrhythmias, and cardiac arrhythmias (Supplemental Figure 3). We excluded individuals using ACEi, vasodilator anti-hypertensives, and centrally acting anti-hypertensives to mitigate potential confounding effects associated with these medications. Analysis in unmatched data was adjusted for sex and age of disease diagnosis, while analysis in matched data was unadjusted.

In this study, we employed the SurvSNP package (PMID: 22685040) to conduct power calculations, leveraging asymptotic estimates derived from genetic hazard ratios, event rates, study population size, and allele frequencies. These calculations were based on Cox proportional hazards model results obtained before and after genetic matching for rs11591147 and rs562556 polymorphisms within the PCSK9 gene. Our objective was to assess the extent of power enhancement achieved through our methodology when examining the time to severe cardiovascular outcomes from the first available diagnosis of IHD. Power calculations for all SNPs can be seen in Supplemental Table 3. Notably, for rs11591147, the power within the unmatched subset was 9.5, demonstrating a substantial increase to 73.4 after matching. Similarly, for rs562556, the power within the unmatched subset was 6.9, exhibiting an increase to 11.16 after matching. These findings underscore the enhancements in statistical power attainable through matched subset analysis, underscoring the pivotal role of selecting appropriate SNPs with substantial effect sizes for robust survival analysis outcomes. (Supplemental Figure 6)

To comprehensively elucidate the diverse array of phenotypic manifestations associated with each of the SNPs, we performed phenome-wide association study (PHEWAS) adjusted for age, sex and 10 genetic principal components, and applied additive model. The PHEWAS analyses were performed using the PHESANT (PHEnome Scan ANalysis Tool) software application (PMID: 29040602). This tool facilitates automated phenome scans within the UK Biobank dataset, thereby enabling comprehensive exploration of phenotypic associations. The main dataset from the UK Biobank downloaded in March 2023 was supplemented with custom cardiovascular phenotypes created using HES update version September 2023. For logistic regression we only included phenotypes with cases > 50.

---

**Supplemental Table 1:**
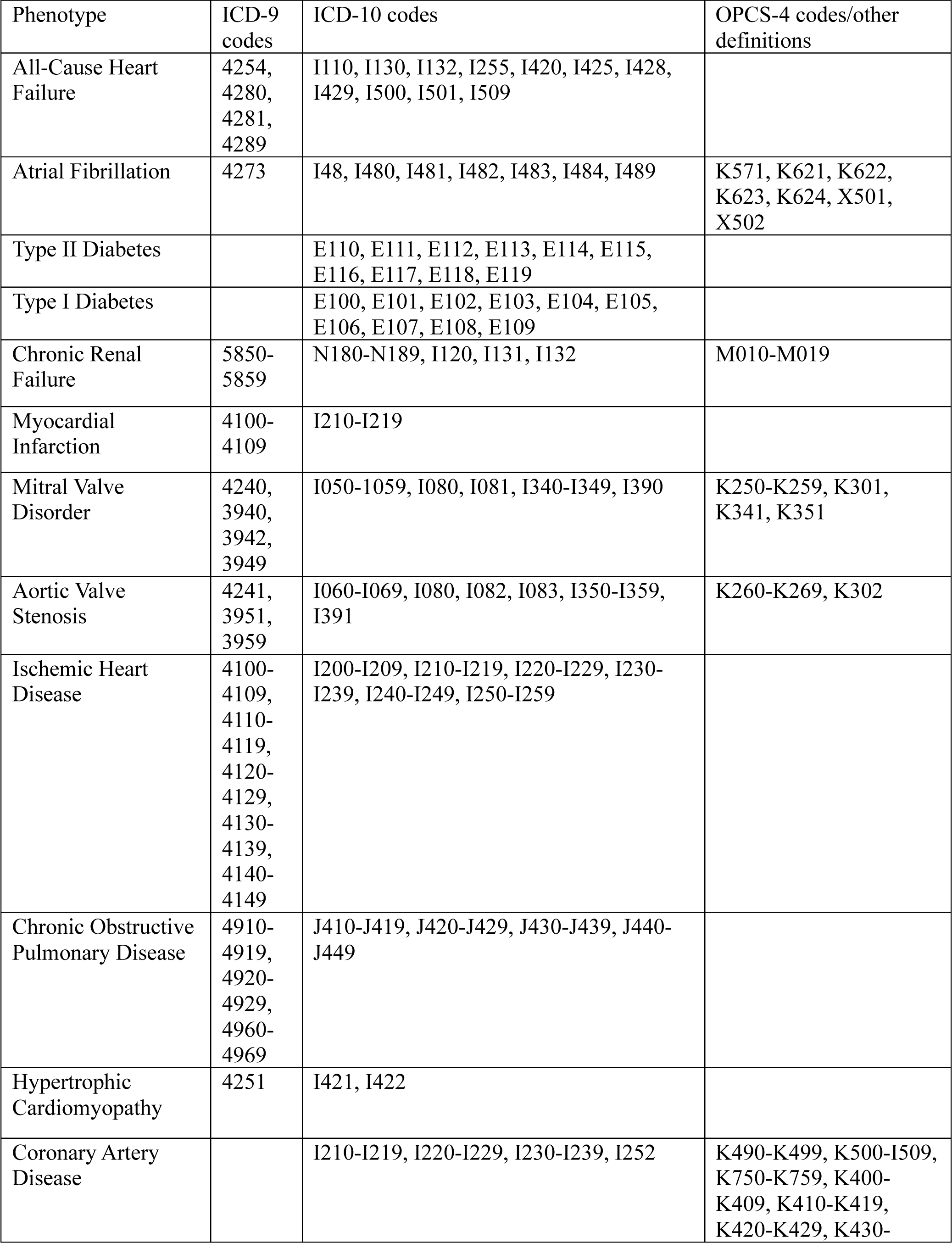
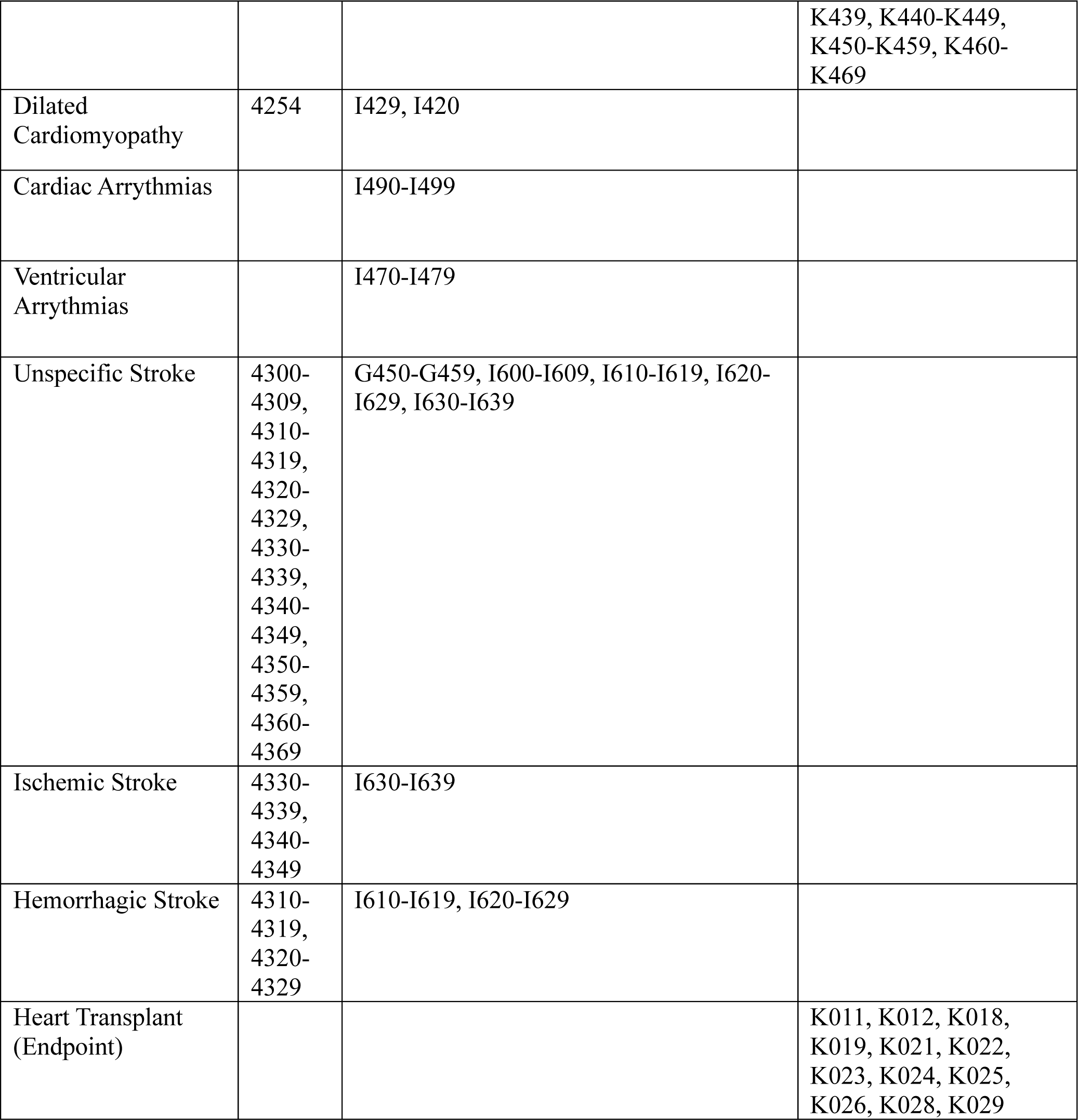
Phenotype definitions for cardiovascular endpoints and comorbidities used for propensity score matching.

**Supplemental Table 2:**
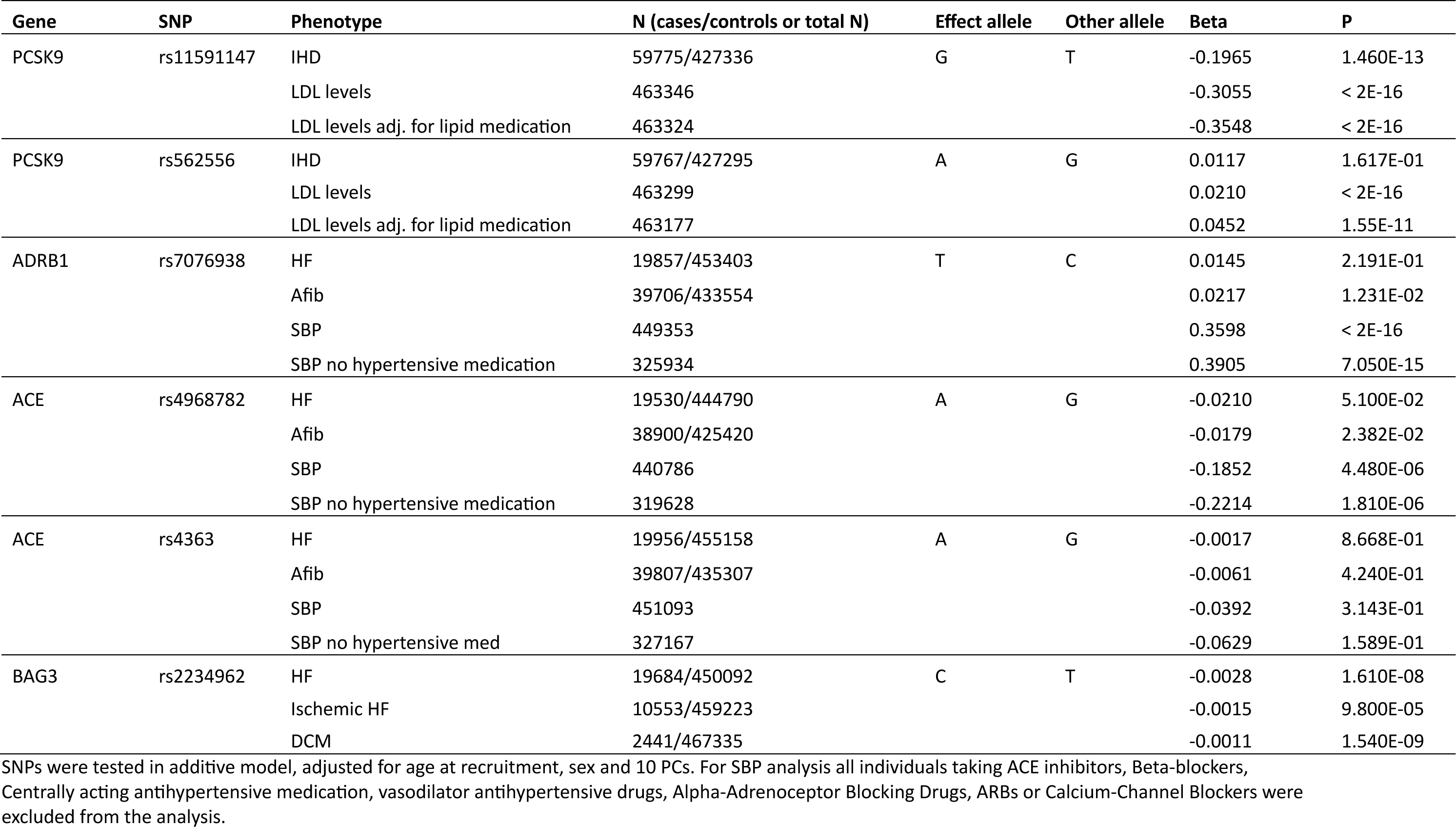
Previously known associations for the tested SNPs.

| Gene | SNP | Phenotype | N (cases/controls or total N) | Effect allele | Other allele | Beta | P |
| --- | --- | --- | --- | --- | --- | --- | --- |
| PCSK9 | rs11591147 | IHD | 59775/427336 | G | T | -0.1965 | 1.460E-13 |
|  |  | LDL levels | 463346 |  |  | -0.3055 | < 2E-16 |
|  |  | LDL levels adj. for lipid medication | 463324 |  |  | -0.3548 | < 2E-16 |
| PCSK9 | rs562556 | IHD | 59767/427295 | A | G | 0.0117 | 1.617E-01 |
|  |  | LDL levels | 463299 |  |  | 0.0210 | < 2E-16 |
|  |  | LDL levels adj. for lipid medication | 463177 |  |  | 0.0452 | 1.55E-11 |
| ADRB1 | rs7076938 | HF | 19857/453403 | T | C | 0.0145 | 2.191E-01 |
|  |  | Afib | 39706/433554 |  |  | 0.0217 | 1.231E-02 |
|  |  | SBP | 449353 |  |  | 0.3598 | < 2E-16 |
|  |  | SBP no hypertensive medication | 325934 |  |  | 0.3905 | 7.050E-15 |
| ACE | rs4968782 | HF | 19530/444790 | A | G | -0.0210 | 5.100E-02 |
|  |  | Afib | 38900/425420 |  |  | -0.0179 | 2.382E-02 |
|  |  | SBP | 440786 |  |  | -0.1852 | 4.480E-06 |
|  |  | SBP no hypertensive medication | 319628 |  |  | -0.2214 | 1.810E-06 |
| ACE | rs4363 | HF | 19956/455158 | A | G | -0.0017 | 8.668E-01 |
|  |  | Afib | 39807/435307 |  |  | -0.0061 | 4.240E-01 |
|  |  | SBP | 451093 |  |  | -0.0392 | 3.143E-01 |
|  |  | SBP no hypertensive med | 327167 |  |  | -0.0629 | 1.589E-01 |
| BAG3 | rs2234962 | HF | 19684/450092 | C | T | -0.0028 | 1.610E-08 |
|  |  | Ischemic HF | 10553/459223 |  |  | -0.0015 | 9.800E-05 |
|  |  | DCM | 2441/467335 |  |  | -0.0011 | 1.540E-09 |

SNPs were tested in additive model, adjusted for age at recruitment, sex and 10 PCs. For SBP analysis all individuals taking ACE inhibitors, Beta-blockers, Centrally acting antihypertensive medication, vasodilator antihypertensive drugs, Alpha-Adrenoceptor Blocking Drugs, ARBs or Calcium-Channel Blockers were excluded from the analysis.

**Supplemental Table 3:** Power calculations for survival analysis.

| **Gene** | **RSNUM** | **Diagnosis** | **Endpoint** | **Population** | **Matched** | **Power (%)** |
| --- | --- | --- | --- | --- | --- | --- |
| PCSK9 | rs11591147 | Ischemic Heart Disease | CVD/Heart Transplant | Caucasians | No | 9.5 |
| PCSK9 | rs11591147 | Ischemic Heart Disease | CVD/Heart Transplant | Caucasians | Yes | 73.4 |
| PCSK9 | rs562556 | Ischemic Heart Disease | CVD/Heart Transplant | Caucasians | No | 6.9 |
| PCSK9 | rs562556 | Ischemic Heart Disease | CVD/Heart Transplant | Caucasians | Yes | 11.16 |
| ACE | rs4968782 | AF | CVD/Heart Transplant | Caucasians | No | 59.23 |
| ACE | rs4968782 | AF | CVD/Heart Transplant | Caucasians | Yes | 80.28 |
| ACE | rs4968782 | HF | HF Composite | All | No | 79.81 |
| ACE | rs4968782 | HF | HF Composite | All | Yes | 92.23 |
| ACE | rs4363 | AF | CVD/Heart Transplant | Caucasians | No | 19.93 |
| ACE | rs4363 | AF | CVD/Heart Transplant | Caucasians | Yes | 22.97 |
| ACE | rs4363 | HF | HF Composite | All | No | 5.65 |
| ACE | rs4363 | HF | HF Composite | All | Yes | 17.96 |
| ADRB1 | rs7076938 | AF | CVD/Heart Transplant | All | No | 98.09 |
| ADRB1 | rs7076938 | AF | CVD/Heart Transplant | All | Yes | 96.15 |
| ADRB1 | rs7076938 | AF | AF Composite | All | No | 8.75 |
| ADRB1 | rs7076938 | AF | AF Composite | All | Yes | 98.4 |
| BAG3 | rs2234962 | HF | HF Composite | All | No | 68.11 |
| BAG3 | rs2234962 | HF | HF Composite | All | Yes | 77.88 |
| BAG3 | rs2234962 | Ischemic HF | HF Composite | All | No | 26.58 |
| BAG3 | rs2234962 | Ischemic HF | HF Composite | All | Yes | 51.89 |

**Supplemental Figure 1.**
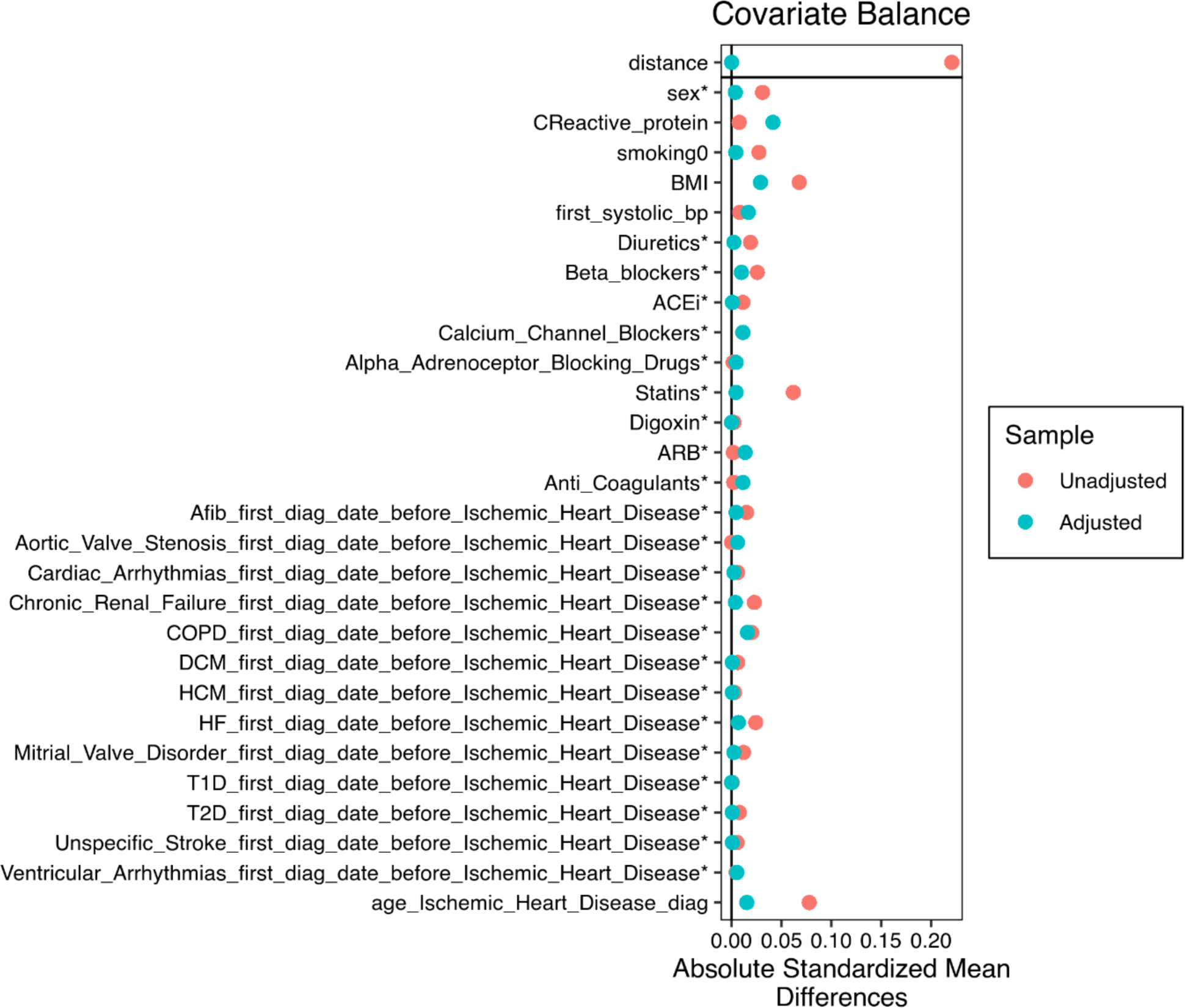
Matching results for PCSK9 11591147 in Caucasians. Love plot indicates shift towards 0 in absolute standardized mean difference, representing successful matching. Asterisk (*) indicates variables for which the displayed value is the raw (unstandardized) difference in means.

**Supplemental Figure 2.**
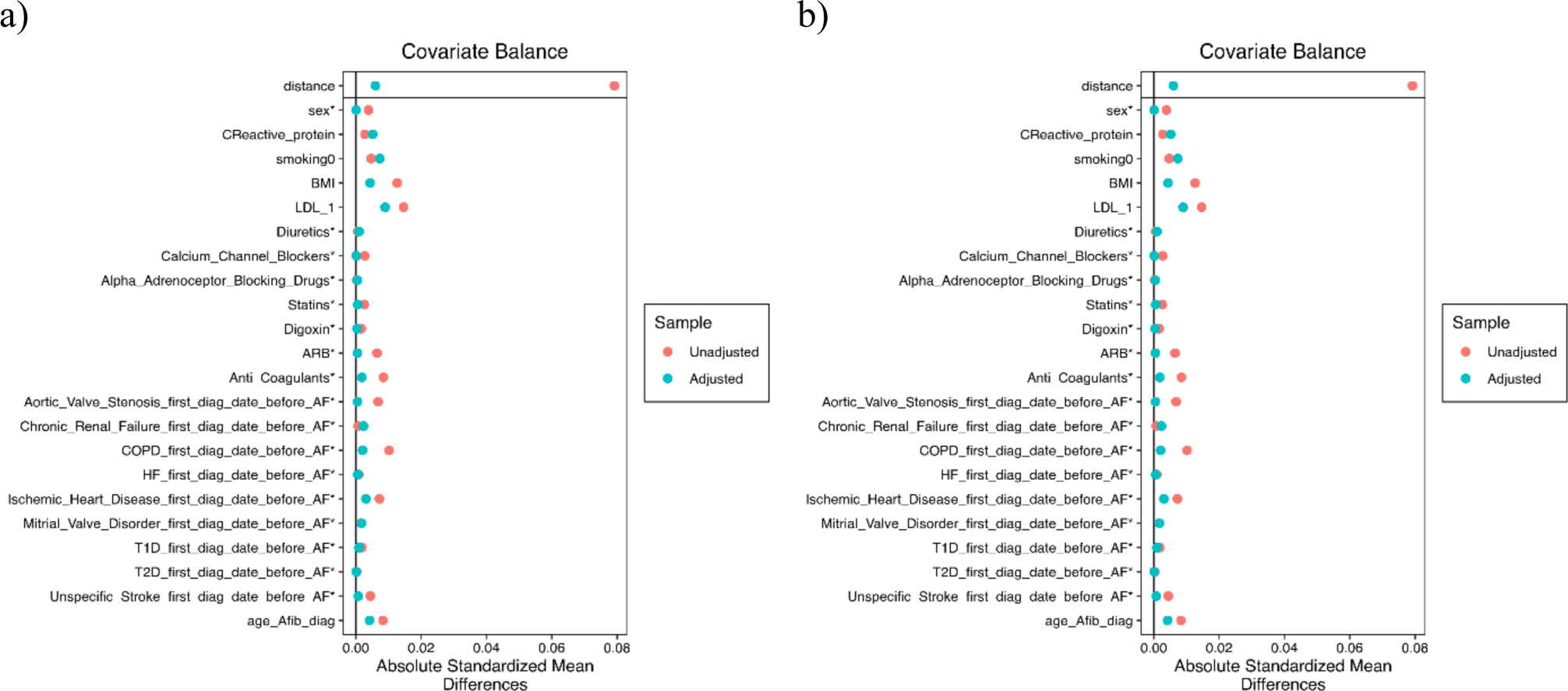
Propensity score matching results for ADRB1 rs7076938 for a) time to cardiovascular (CV) death/heart transplant from first atrial fibrillation diagnosis (AF), and b) time to rehospitalization or CV death/heart transplant due to AF. A shift towards 0 for absolute standardized mean difference indicates improvement in differences between populations carrying specific alleles.

**Supplemental Figure 3.**
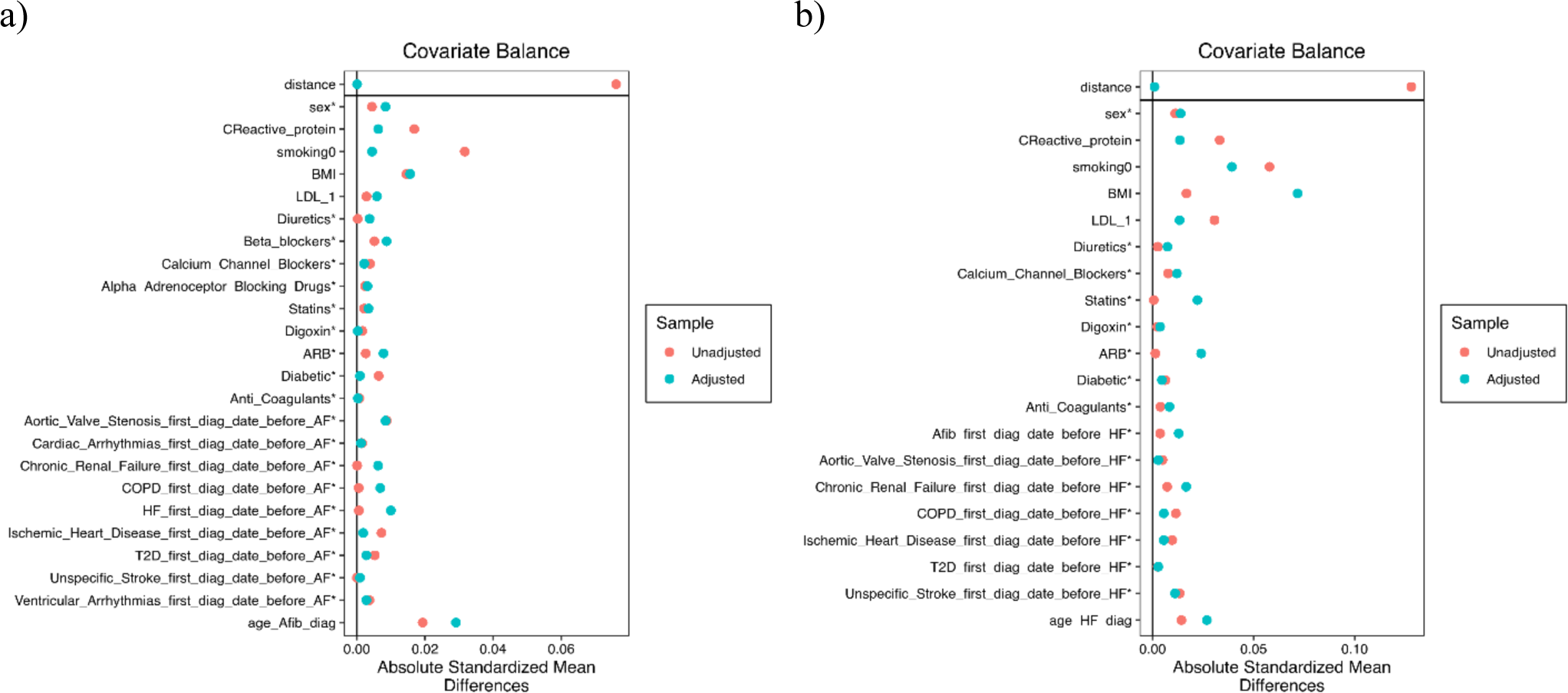
Propensity score matching results for ACE rs4968782 for a) time to cardiovascular (CV) death/heart transplant from first atrial fibrillation (AF) diagnosis in Caucasians, and b) time to rehospitalization or CV death/heart transplant due to all-cause heart failure (HF) in all ethnicities. A shift towards 0 for absolute standardized mean difference represents successful matching.

**Supplemental Figure 4.**
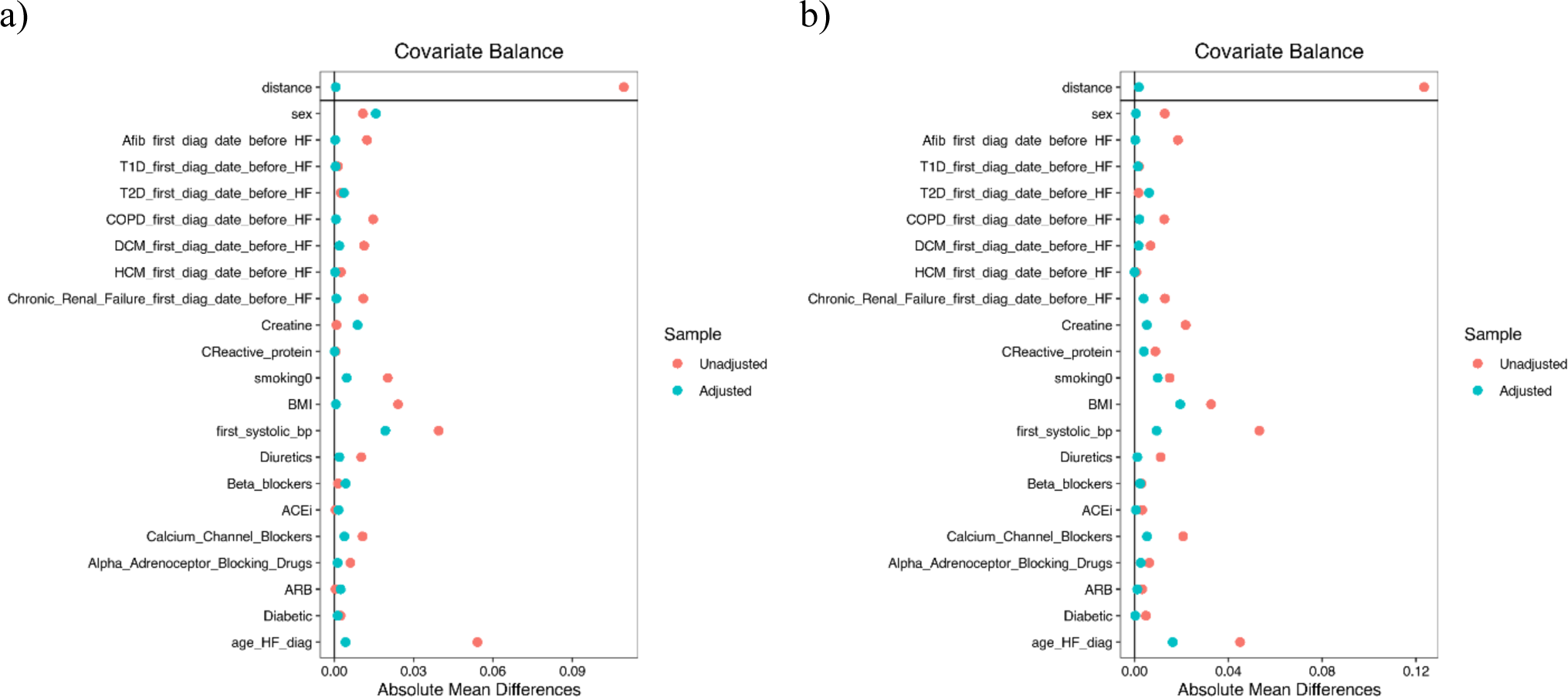
Propensity score matching results for BAG3 for a) all-cause HF patients, and b) Ischemic HF patients. A shift towards 0 for absolute standardized mean difference represents successful matching.

**Supplemental Figure 5.**
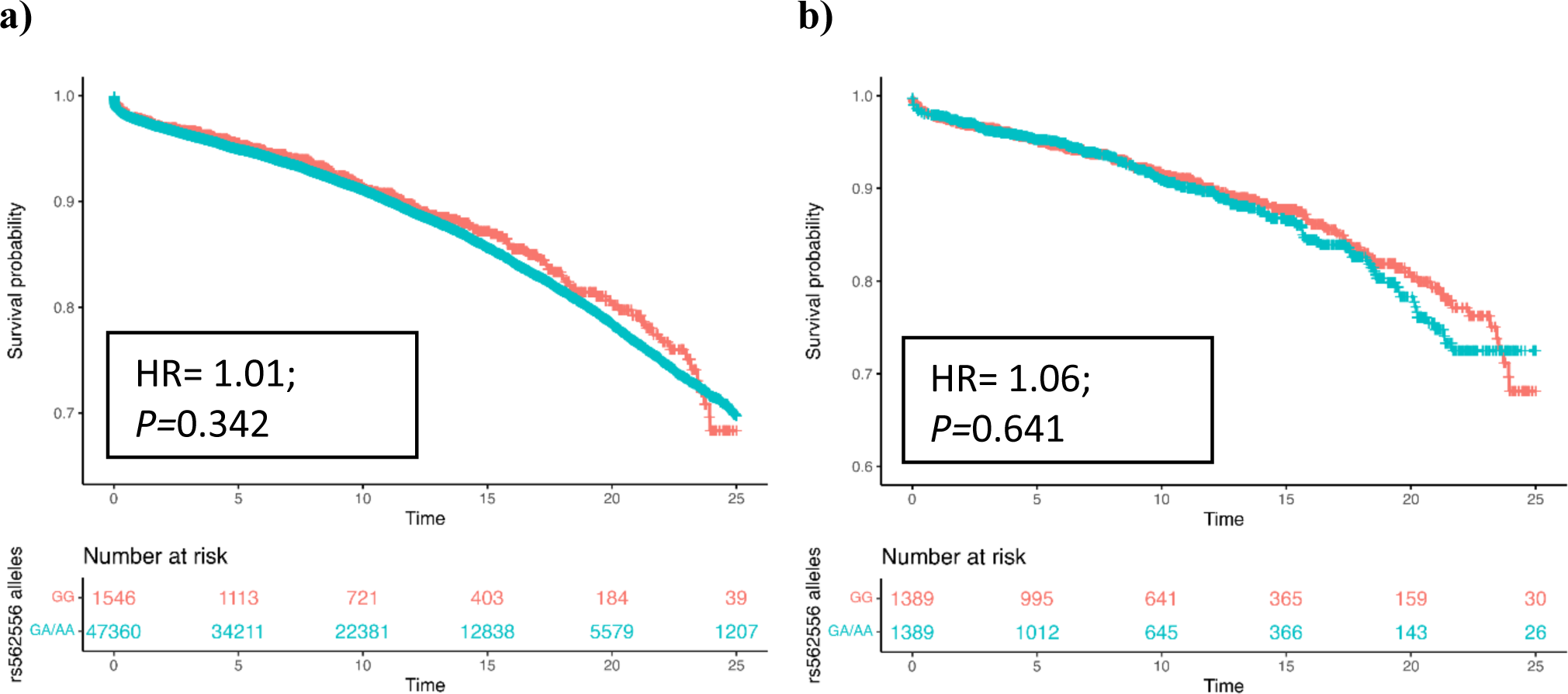
Effect of PCSK9 rs562556 on time to cardiovascular (CV) death/heart transplant from first ischemic heart disease (IHD) diagnosis in a) unmatched and b) matched participants. Participants were filtered for Caucasian ethnicity. The time variable represents years to outcome while the allele variable represents control and treatment groups (those with GG genotype versus those with GA/AA genotype).

**Supplemental Figure 6.**
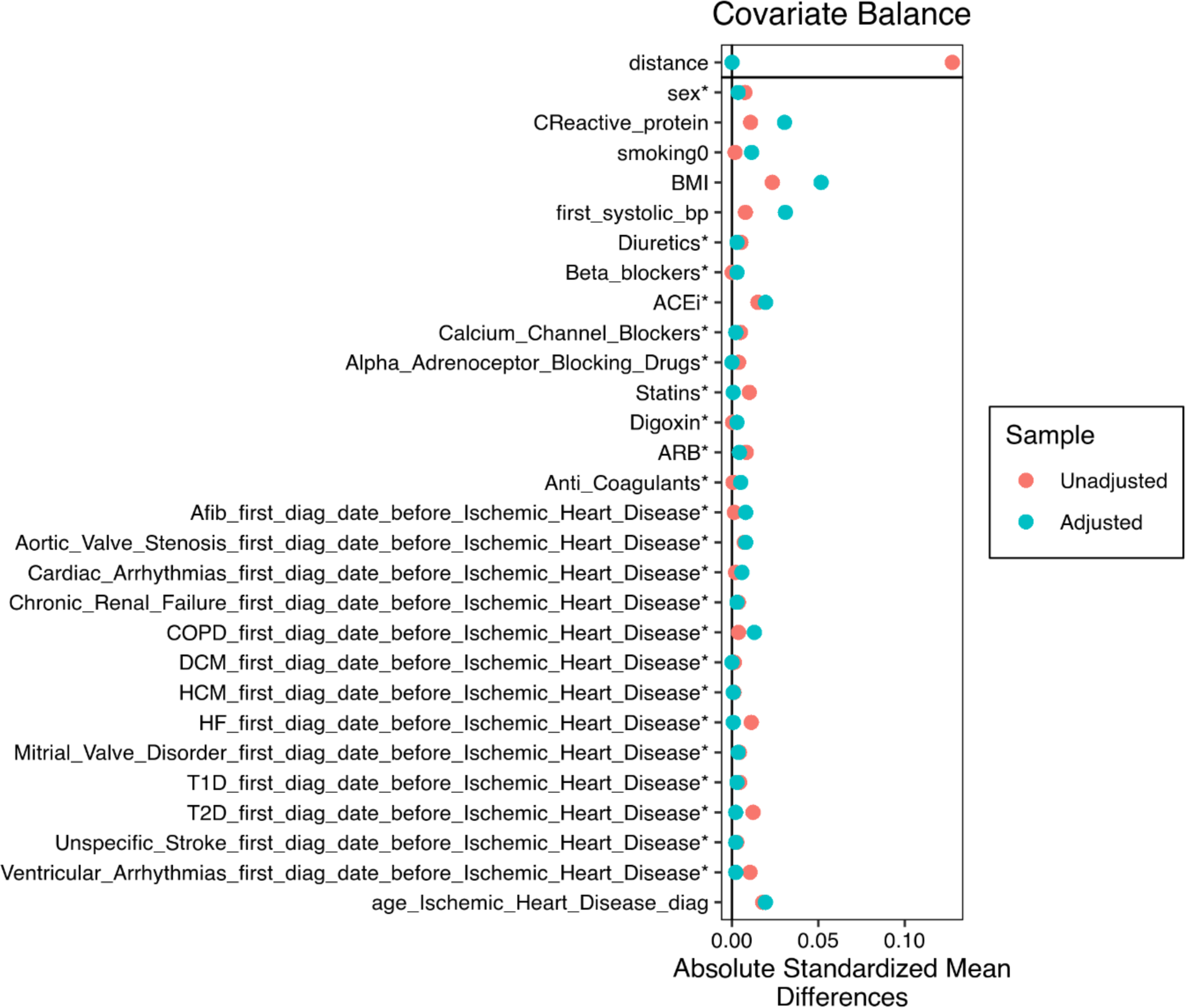
Matching results for PCSK9 rs562556 in Caucasians. Shift towards 0 in absolute standardized mean difference indicates successful matching. Asterisk (*) indicates variables for which the displayed value is the raw (unstandardized) difference in means.

**Supplemental Figure 7.**
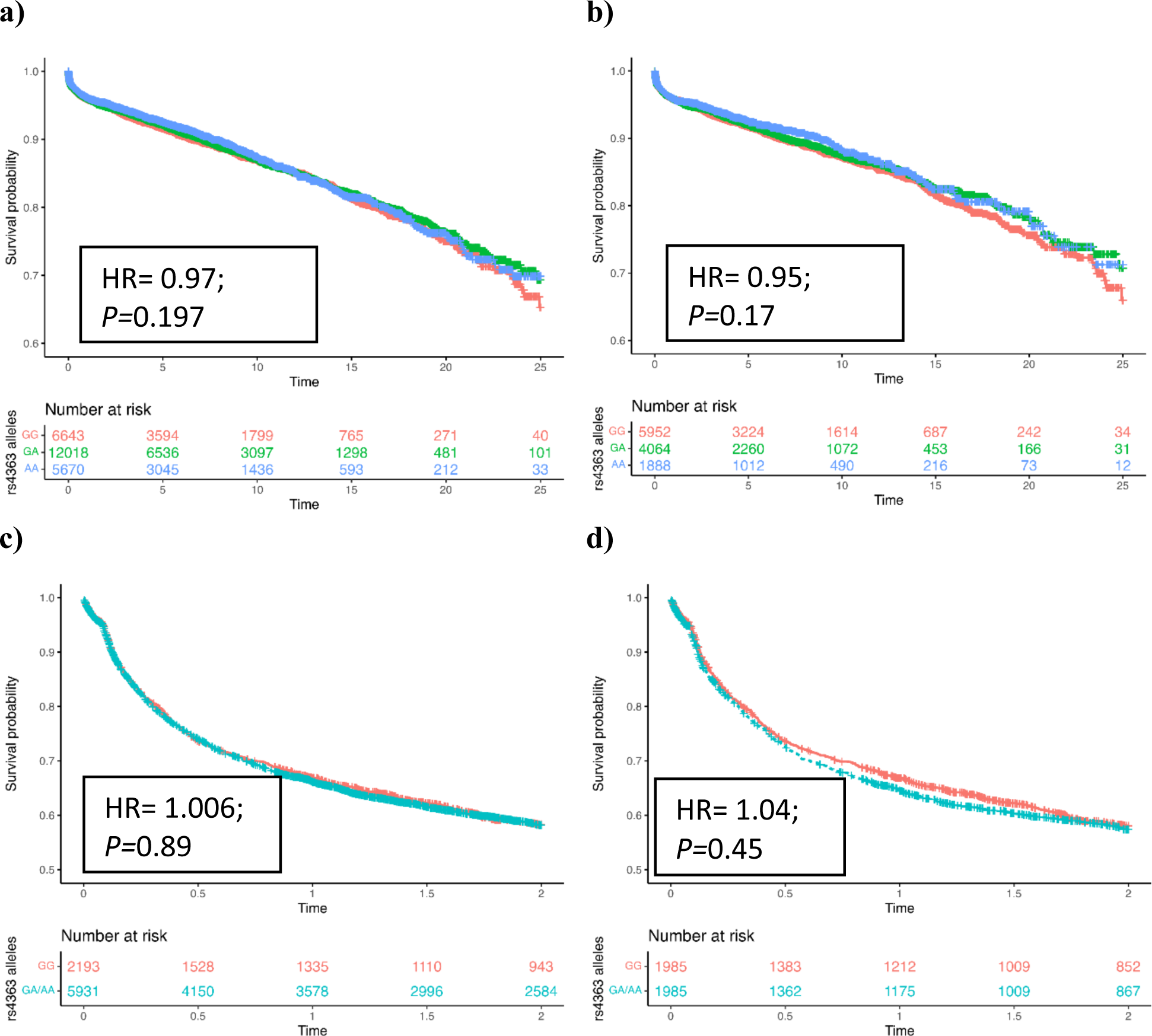
Effect of ACE rs4363 on time to cardiovascular (CV) death/heart transplant from first atrial fibrillation (AF) diagnosis in a) unmatched and b) matched Caucasian participants, and time to rehospitalization or CV death/heart transplant due to all cause heart failure (HF) in c) unmatched and d) matched (all ethnicities) participants. The time variable represents years to severe cardiovascular outcome (death or transplant) while the allele variable represents control and treatment groups (GG genotype versus GA/AA genotype).

**Supplemental Figure 8.**
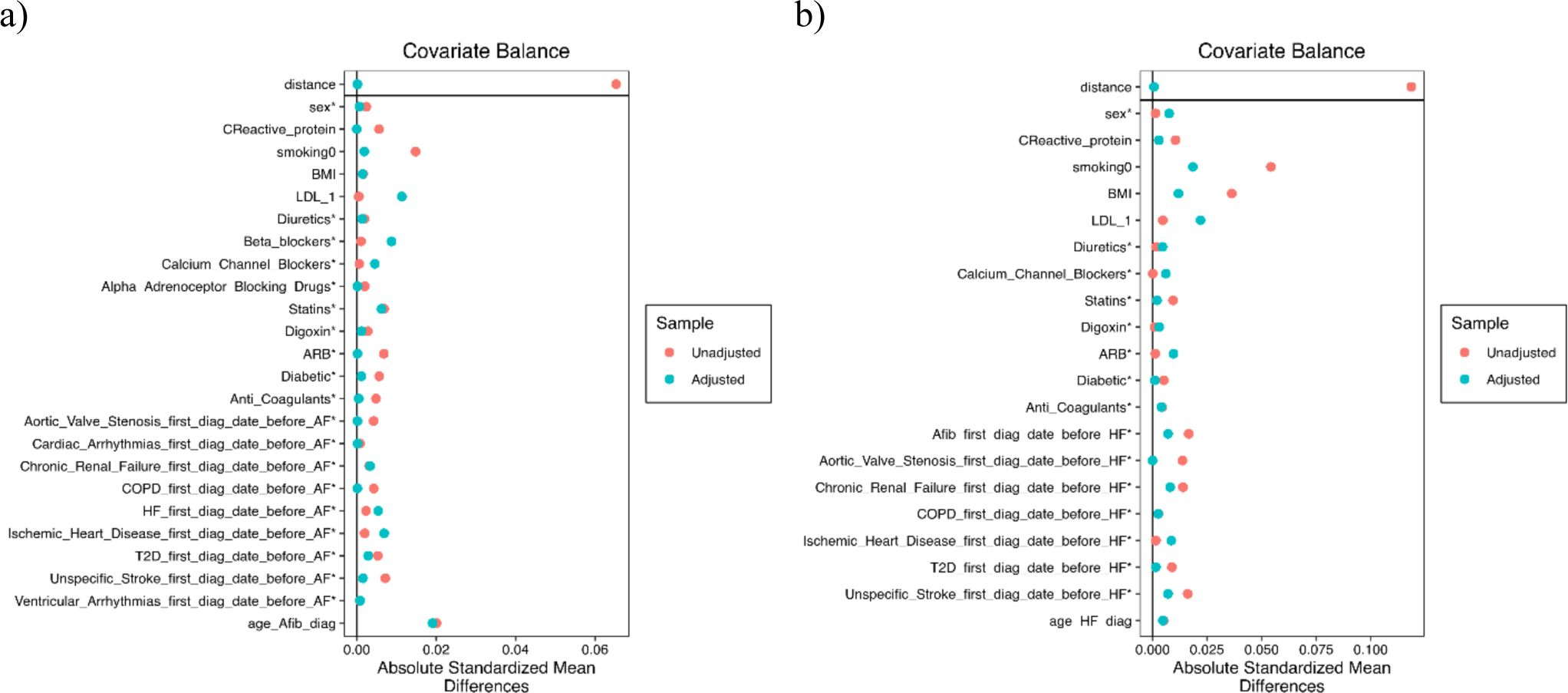
Propensity score matching results for ACE rs4363 for a) time to cardiovascular (CV) death/heart transplant from first atrial fibrillation (AF) diagnosis in Caucasians, and b) time to rehospitalization or CV death/heart transplant due to all-cause heart failure (HF) in all ethnicities. A shift towards 0 for absolute standardized mean difference represents successful matching.

**Supplemental Figure 9.**
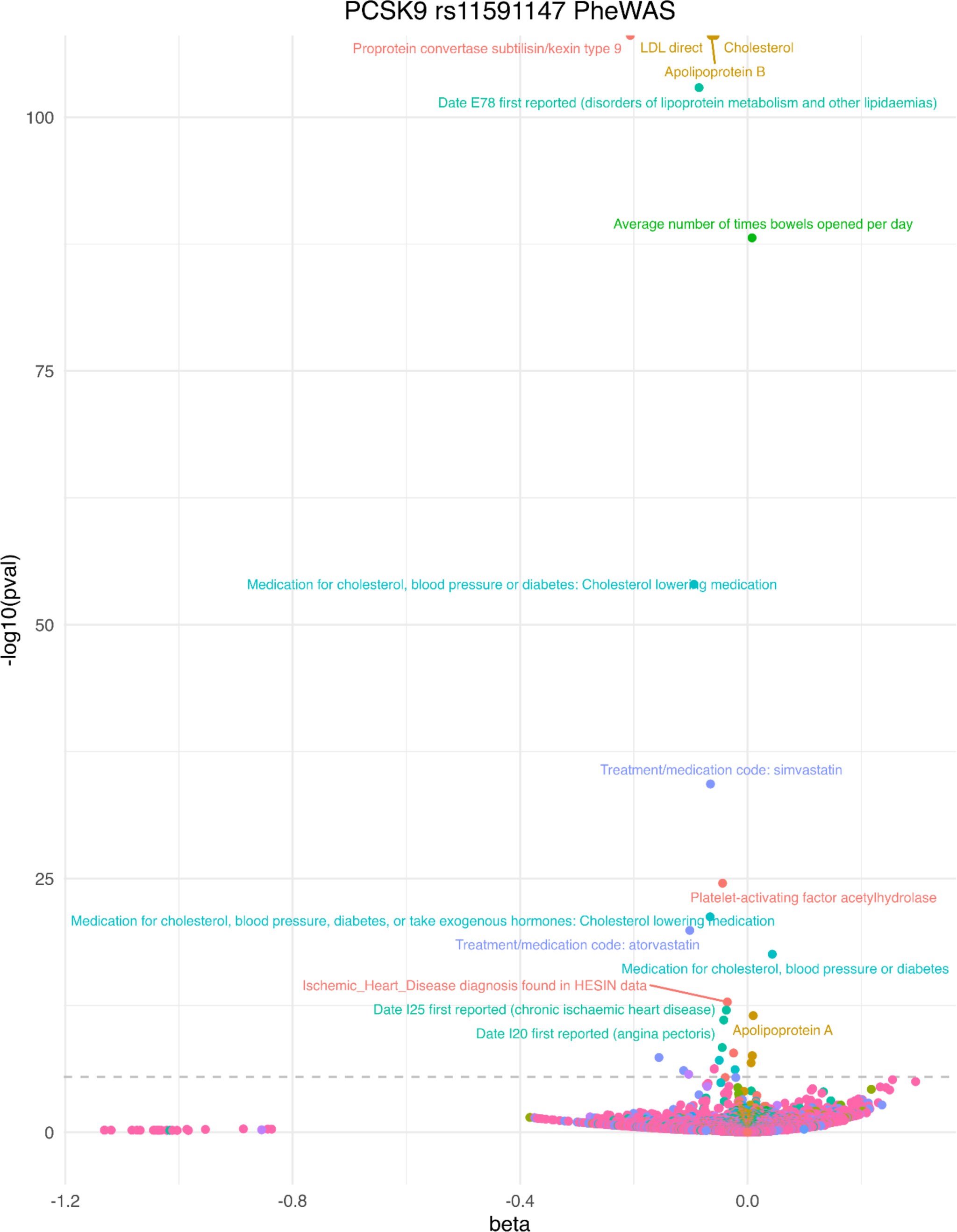
PheWAS results for PCSK9 rs11591147 which includes all participants with available genotype data and adjusted for sex, age of recruitment, and ten genetic principal components.

**Supplemental Figure 10.**
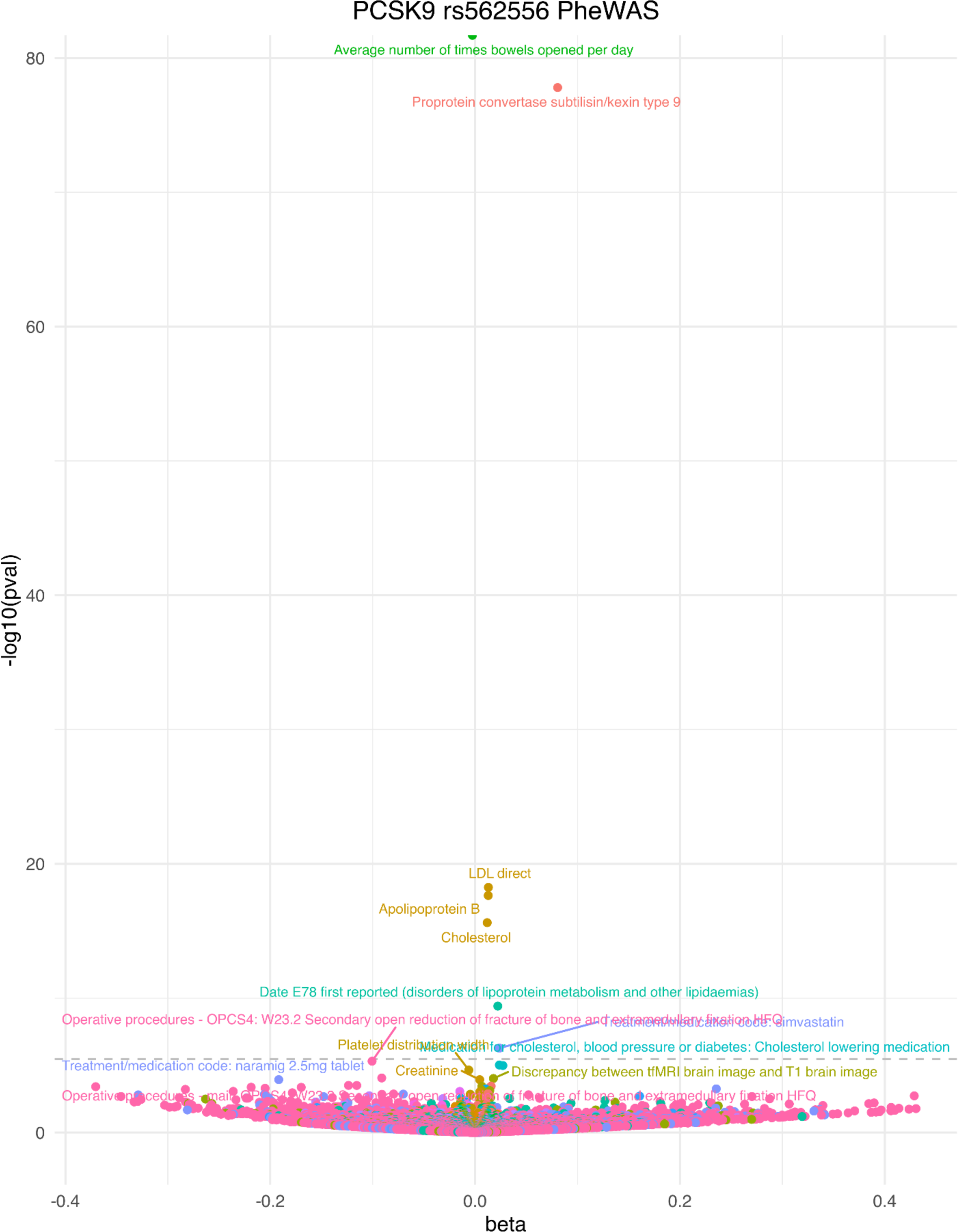
PheWAS results for PCSK9 rs562556. Analysis for all participants with available genotype data, adjusted for sex, age of recruitment, and 10 genetic principal components.

**Supplemental Figure 11.**
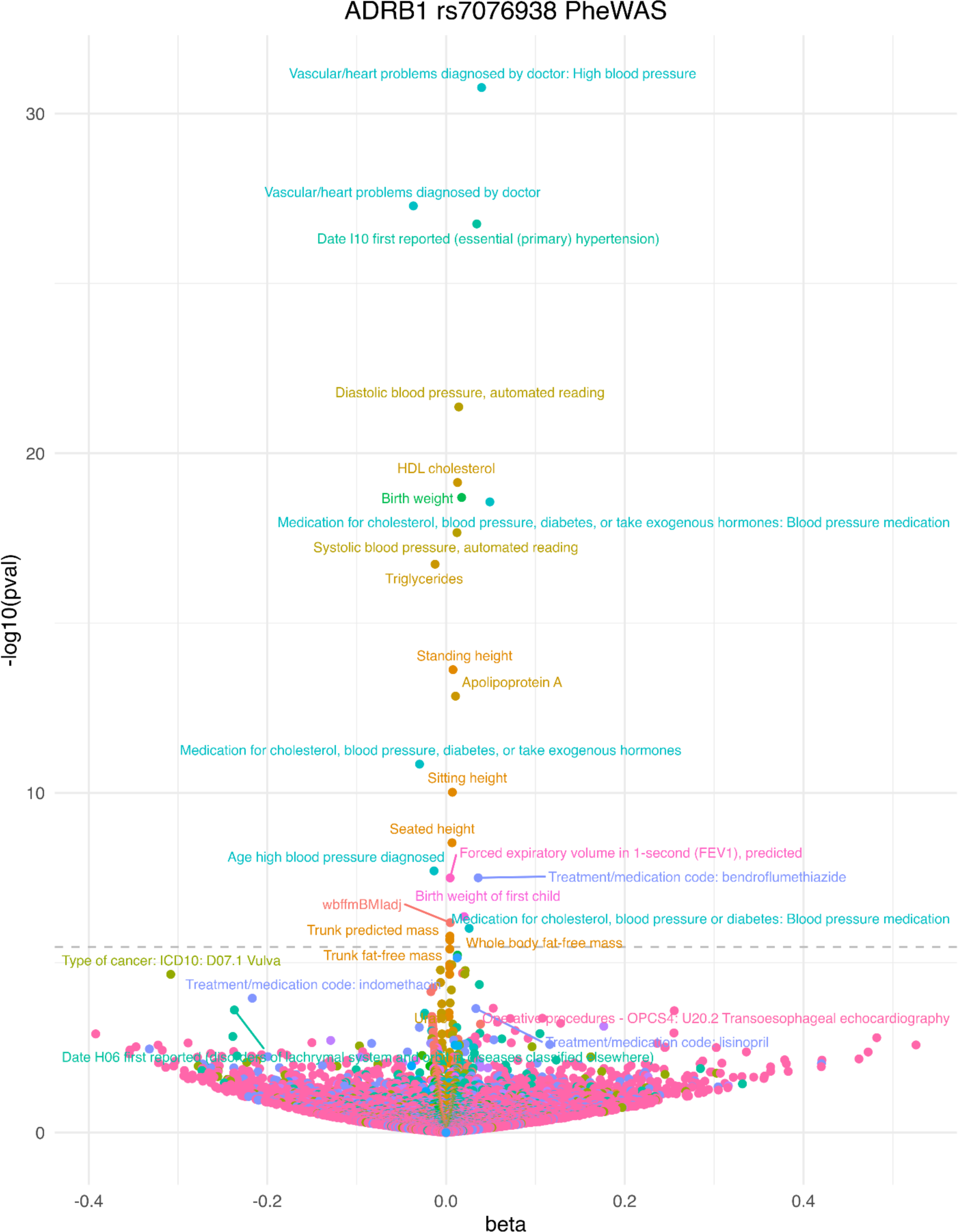
PheWAS results for ADRB1 rs7076938 (X allele). Analysis for all participants with available genotype data, adjusted for sex, age of recruitment, and 10 genetic principal components.

**Supplemental Figure 12.**
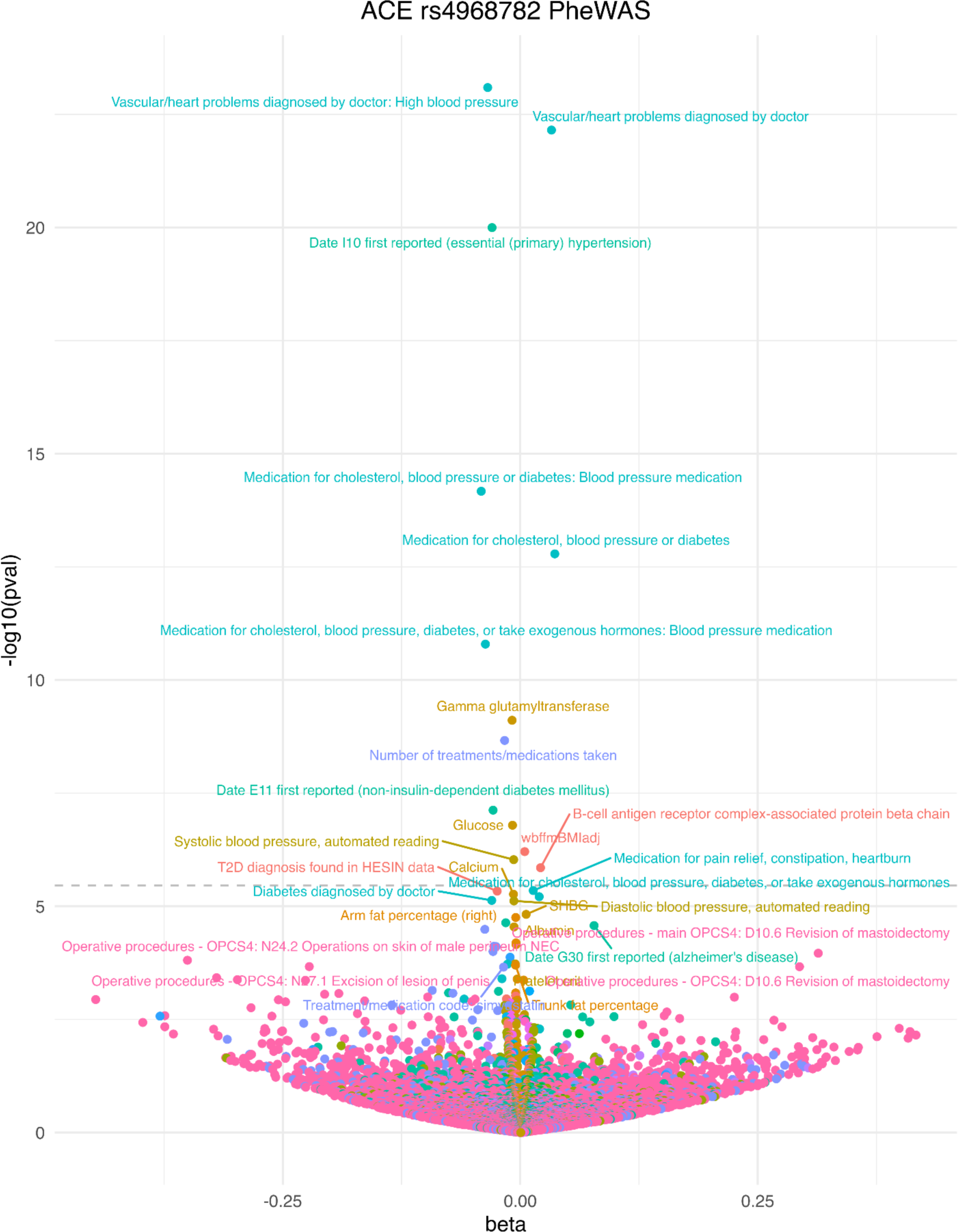
PheWAS results for ACE rs4968782. Analysis for all participants with available genotype data, adjusted for sex, age of recruitment, and 10 genetic principal components.

**Supplemental Figure 13.**
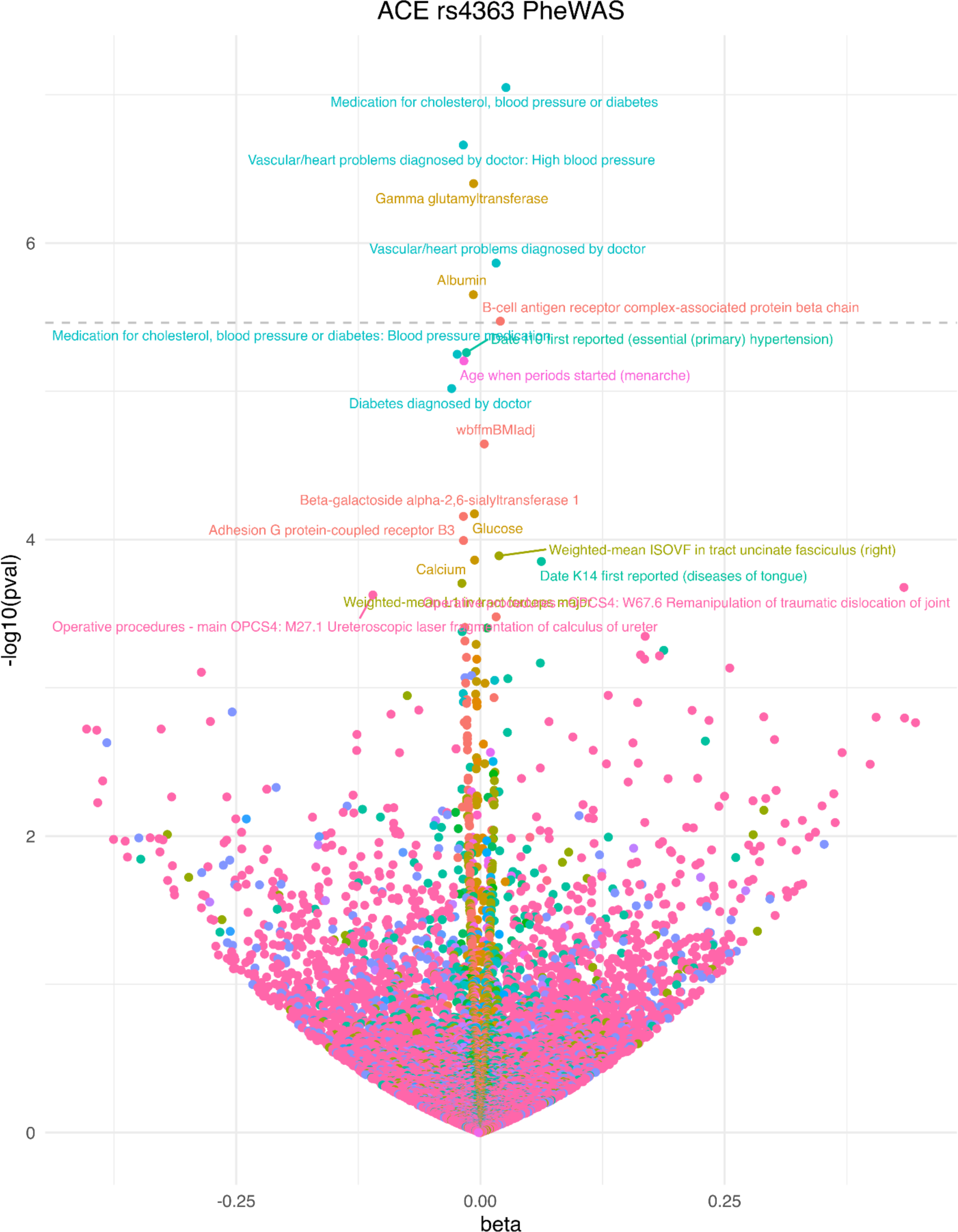
PheWAS results for ACE rs4363. Analysis for all participants with available genotype data, adjusted for sex, age of recruitment, and 10 genetic principal components.

**Supplemental Figure 14.**
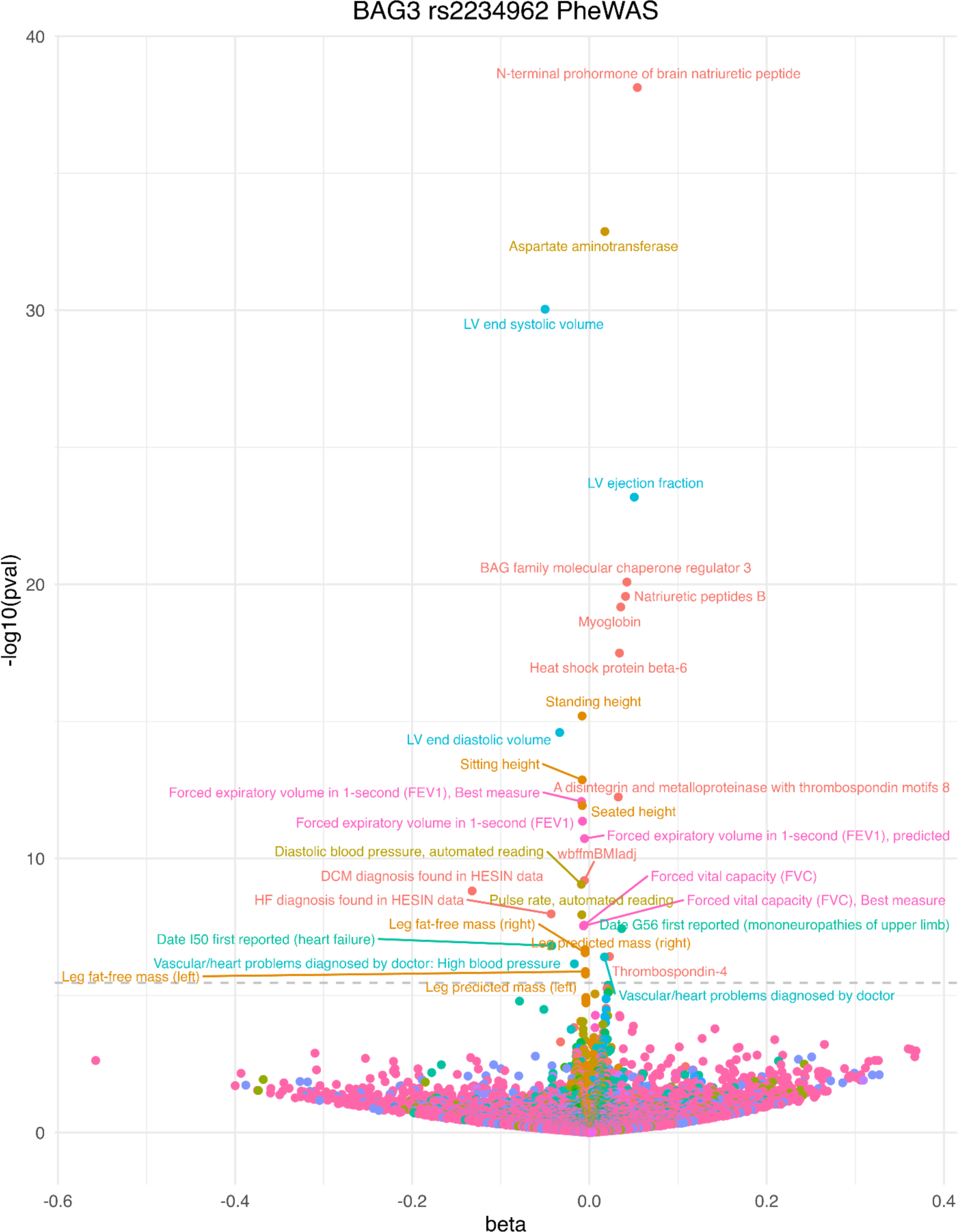
PheWAS results for BAG3 rs2234962. Analysis for all participants with available genotype data, adjusted for sex, age of recruitment, and 10 genetic principal components.

## Notes

### Competing Interest Statement

All authors are employees or contractors of Tenaya Therapeutics a publicly traded biopharmaceutical company exclusively focused on Cardiovascular Disease. FF, WY, TH, WW JRP, SMF are shareholders of Tenaya Therapeutics

### Funding Statement

This study was performed by employees and contractors of Tenaya Therapeutics

### Author Declarations

Genetic and clinical data in the UKB cohort were obtained from the UKB (https://www.ukbiobank.ac.uk) and is available to researchers through a streamlined application process. The UK Biobank was approved by the North West Multi-Centre Research Ethics Committee and all participants provided written informed consent to participate in the study.

